# Nasal foralumab treatment of PIRA induces regulatory immunity, dampens microglial activation and stabilizes clinical progression in non-active secondary progressive MS

**DOI:** 10.1101/2025.04.30.25326602

**Authors:** Tanuja Chitnis, Tarun Singhal, Jonathan Zurawski, Taylor J. Saraceno, Niveditha Gopalakrishnan, Lauren Cain, Brenna LaBarre, Devin King, Regan W. Bergmark, Alice Z. Maxfield, Steven Cicero, Hong Pan, Shipra Dubey, Steven Vaquerano, Caleb Hansel, Brian Healy, Shrishti Saxena, Hrishikesh Lokhande, Moogeh Baharnoori, Evan Madill, Manali Sheth, Rachel Rodin, Jessica Ye, Nancy Clementi, William A. Clementi, Howard L. Weiner

**Affiliations:** Harvard Medical School, Boston, MA; Ann Romney Center for Neurologic Diseases, Brigham and Women’s Hospital, Mass General Brigham, Boston, MA; Brigham Multiple Sclerosis Center, Department of Neurology, Brigham and Women’s Hospital, Mass General Brigham, Boston, MA; Division of Nuclear Medicine, Department of Radiology, Brigham and Women’s Hospital, Mass General Brigham, Boston, MA; Department of Otolaryngology, Brigham and Women’s Hospital, Mass General Brigham, Boston, MA; Clementi Associates, Bryn Mawr PA; Tiziana Life Sciences, New York, NY

## Abstract

**Background:** Progression independent of relapses (PIRA) is a major therapeutic challenge in multiple sclerosis (MS). Nasal anti-CD3 treats animal models of progressive MS by inducing regulatory T cells (Tregs) that suppress central nervous system (CNS) inflammation and lessen clinical disease.

**Methods:** Ten patients with non-active secondary progressive MS (naSPMS) that continued to progress on B cell therapy were treated with nasal anti-CD3 (foralumab) for a minimum of six months in an open label study. Safety monitoring included otolaryngology evaluation and neurologic assessments including Expanded Disability Status Scale (EDSS), Multiple Sclerosis Functional Composite (MSFC-4), Modified Fatigue Impact Scale (MFIS), California Verbal Learning Test (CVLT-II) and Low Contract Visual Acuity (LCVA). MRI and microglial translocator protein (TSPO)-PET imaging with [F-18]PBR06 were conducted. Serum and cerebrospinal fluid (CSF) proteomic biomarkers and single cell RNA sequencing of blood was performed to evaluate foralumab-induced immunomodulation. The endpoints of our study were safety, clinical effects, microglial signal and immune measures.

**Results:** All patients stabilized on EDSS scores and three of four patients treated continuously for 12 months had improvement on EDSS. Six of 10 patients had improvement in fatigue on the MFIS scale. There were no treatment-related serious adverse events (SAEs) or severe AEs and no new T2 lesions were observed on MRI. There was a reduction in TSPO-PET signal over six months (p<0.05). Changes in peripheral blood gene expression occurred as early as three months and affected antigen presentation, interferon responses and regulatory pathways in multiple cell types including FoxP3+ Tregs, CD4+ Tcm cells, CD8+ Tem cells, CD14+ and CD16+ monocytes and B cells. TGFβ expression was increased across cell multiple subsets.

**Interpretation:** These findings identify a novel, non-toxic immune based therapy for the treatment for PIRA that acts by the induction of a regulatory immune responses and dampens microglial inflammation. Double blind placebo-controlled trials are warranted to explore nasal foralumab for the treatment of naSPMS.

## INTRODUCTION

Progression independent of relapses (PIRA) is a major therapeutic challenge in multiple sclerosis (MS).^1, 2^ Current MS treatments have a beneficial effect on the relapsing-inflammatory component of MS but have limited efficacy on PIRA. PIRA and progressive forms of MS are characterized by microglial activation and dysfunctional astrocytes^3–6^ which result in ongoing central nervous system (CNS)-centric inflammation and neurodegeneration. Current therapeutic approaches in MS that largely consist of B cell depletion or inhibition of cell trafficking with sphingosine-1-phosphate modulators or alpha-4-integrin antibodies, have a limited effect on CNS-centric or trapped inflammation behind the blood-brain-barrier. Novel approaches to suppress CNS-centric inflammation in progressive forms of MS are urgently needed.^6^

Mucosal surfaces, such as the gastrointestinal tract and the nasal cavity, have a well-developed immune system by which the organism interfaces with the environment and which play a key role in immune homeostasis.^7^ Immune interactions at mucosal surfaces induce tolerogenic immune responses, of which regulatory T cells (Treg) are a major component.^8^ The induction of Tregs at mucosal surfaces by oral or nasal antigen treats a large variety of autoimmune and inflammatory diseases in animal models.^9^ In addition, we found that mucosal administration of anti-CD3 monoclonal antibody (mAb) also treats autoimmune and inflammatory models by binding to the T cell receptor and inducing Tregs.^10, 11^ Intravenous anti-CD3 mAb has been used to treat disease beginning with OKT3/muromab to treat transplant rejection (reviewed in Kuhn et al^12^) and more recently with teplizumab to treat type 1 diabetes.^13^

Given the importance of progressive MS, we tested both oral and nasal anti-CD3 mAb in the non-obese diabetic (NOD) progressive EAE model. Although oral anti-CD3 treated acute EAE,^11^ we found that in progressive EAE nasal (but not oral) anti-CD3 attenuated disability, reduced demyelination and axonal damage and dampened microglia and astrocyte activation.^14^ Nasal anti-CD3 acted by inducing LAP+ and IL-10+ Tregs. Nasal anti-CD3 did not enter the brain but localized in the cervical lymph nodes, a key immunogenic organ in the pathogenesis of MS.

In order to translate our findings to humans, we performed a safety and dose-ranging study of foralumab, a fully human anti-CD3 antibody, given nasally to healthy controls for 5 consecutive days at doses of 10ug, 50ug, and 250ug.^15^ We found all three doses to be safe and the 50ug dose to be the most immunomodulatory by dampening effector CD8+ T cell function and inducing a regulatory T cell response. Based on these results, we initiated an open label study of nasal foralumab in naSPMS under the FDA expanded access program. The goals of our study were to assess safety, clinical effects, microglial activation and immune effects following nasal foralumab. As part of safety, patients underwent physical and neurological exams and nasal exams by an ear nose and throat (ENT) specialist. To assess immune effects of nasal foralumab, serum and CSF proteomic biomarkers and blood single cell RNA sequencing was employed.

To evaluate effects of nasal anti-CD3 on microglia, patients underwent translocator protein (TSPO)-PET scanning using [F-18]PBR06 which has been used to study microglia in animals^16^ and humans.^17^ In MS, the TSPO-PET signal predominantly arises from microglia, with some contribution from astrocytes.^18^ In humans, the signal indicates glial density as opposed to a glial activation state.^19^ However, given that TSPO expression is increased in disease-associated microglia in humans,^20^ the TSPO-PET signal is linked to neurodegenerative diseases and thus can serve as a measure of disease and response to therapy. Microglia have been studied extensively in MS using TSPO-PET^21, 22^ and have shown an association of increased TSPO expression in the substantia nigra with fatigue^23^ prediction of PIRA based on increased TSPO signal in normal-appearing white matter, perilesional white matter and thalamus,^24, 25^ higher expression of TSPO in the grey matter and normal appearing white matter in progressive vs relapsing MS,^26, 27^ correlation of TSPO expression in the grey matter with EDSS^26^ and widespread glial activation in primary progressive MS.^28^

Our study demonstrated that nasal foralumab was safe, induced a regulatory immune response, dampened microglial activation and stabilized clinical progression in subjects with naSPMS.

## METHODS

### Patient eligibility

Patients followed at the Brigham MS Center with naSPMS defined by the absence of relapses or new MRI lesions in the past two years were included in this program. Patients were referred by their primary neurologist, and included if they met eligibility criteria, Key inclusion criteria: 1) confirmed diagnosis of MS and MRI consistent with a diagnosis of MS; 2) age 25-70; 3) Failed standard of care treatment and continued to decline clinically for at least 6 months. Key exclusion criteria: 1) corticosteroid treatment in the past 30 days; 2) treatment with Ocrevus, Rituxan, Kesimpta or Truxima in the past 90 days; 3) inability to tolerate nasally administered medications. A full listing of the eligibility criteria is included in the supplementary materials. Additionally, patients were required to be high or medium affinity TSPO-binders evaluated by DNA polymorphism testing. All patients provided informed consent. The protocol was approved by the Mass General Brigham (MGB) IRB. Investigational New Drug Application (IND) approval was obtained for use of foralumab in naSPMS in an expanded access program. The first patient EA1 initiated treatment in May 2021, and following demonstration of safety and efficacy as defined by decreased microglial TSPO-PET signal, additional INDs were approved for EA2, and then EA3-6, followed by EA7-11 (IND 161148). Data obtained between the start of the first patient in May 2021 through September 2024 are presented here.

### Foralumab

Foralumab (TZLS-401, NI-0401) was provided by Tiziana Life Sciences. Foralumab is a fully human IgG1 mAb directed against the CD3ε chain expressed on T cells with a molecular weight of 148,619 Da. Foralumab was administered three times a week (typically, Monday, Wednesday, and Friday) at a total dose of 50µg per day (25µg/100µl to each nostril) for two weeks, followed by a one-week drug holiday. This three-week cycle was repeated for up to six months in some patients and longer in other patients who wished to continue treatment and with the recommendation of their primary treating neurologist. The dose of 50ug was based on our studies in healthy volunteers,^15^ and the dosing regimen was based on immune responses observed in animal preclinical studies. Foralumab was administered using a Gerresheimer syringe (Gerresheimer AG, Vineland, NJ, US) and a VAX300 (TeleFlex, Morrisville, SC, US) nosepiece was placed at the time of drug administration. Initially all doses were administered at the Brigham and Women’s Hospital. In 2022, approval from the FDA and MGB IRB allowed for at-home dosing for all doses excluding the first dose of the three-week cycle.

### Clinical visits

Clinical visits at the BWH took place every three weeks with a physical examination, neurological examination and monitoring bloodwork. Multiple Sclerosis Functional Composite (MSFC-4), Modified Fatigue Impact Scale (MFIS) and California Verbal Learning Test (CVLT-II) were administered every three weeks. A short Quality of Life in Neurological Disorders (NeuroQoL) questionnaire was administered at baseline and every three months. A full schedule of events (SOE) table is provided in the Supplementary materials. Expanded Disability Status Scale (EDSS) scoring was conducted by Neurostatus-certified neurologists.

### Nasal questionnaire and nasal examination

Nasal examinations with nasal endoscope by a board-certified ear, nose, and throat (ENT) physician were conducted at baseline and every three months at minimum. The Nasal Symptoms Questionnaire (NSQ) was administered every three weeks at the first visit of the cycle both prior to dosing and following dosing and once during the rest week.

### Anti-drug antibody assay

Anti-drug antibody (ADA) assay was conducted at Frontage laboratories using the Frontage bioanalytical assay (Frontage Laboratories, Exton, PA, US) in samples from EA1 and EA2 (baseline, three months, four to five months, nine months, 12 months and 15-17 months, as well as in EA3-6 (baseline, three months and six months). An electrochemiluminescence (ECL) immunoassay method (BTM-2761-R2) was developed and validated at Frontage Laboratories, Inc. for the detection (screening, confirmation, and titration) of anti-foralumab antibodies (ADAs) in human serum samples. The screening portion of the method is used to detect potentially positive anti-drug antibodies to foralumab in human serum. The confirmation assay is used to determine if potentially positive samples are specific to foralumab. The titration assay provides a quasi-quantitative estimate of ADA in the positive human serum samples.

### PET genotyping

Participants were genotyped for a polymorphism within the TSPO gene on chromosome 22q13.2, using a Taqman assay; only high and medium affinity binders were included in the study.

<u>PET Acquisition and Analysis</u>: 18F-PBR06 was produced in the PET radiochemistry facility at our hospital and at Perceptive (formerly InVicro, New Haven, CT, US) according to standardized procedures. [F-18]PBR06 was injected as a bolus via an IV catheter in an upper extremity vein; images were acquired in a list acquisition mode using a high-resolution, whole-body PET/CT scanner. Summed PET SUV images based on data acquired between 60 and 90 minutes after tracer injection was co-registered to each individual patient’s T1 MRI (the spin-echo or MPRAGE) scans using the PMOD 3.8/3.9 platform (PMOD Technologies, Zurich, Switzerland; www.pmod.com). This standardized algorithm involved co-registration of the T1-weighted series and PET images of each patient with the Automated Anatomical Template. 60–90-minute SUV images for EA1 were initially evaluated visually. Subsequently, we used z-score based approaches for parametric image generation to evaluate regional changes in the brain as with our previous reports.^28, 29^ Individualized parametric 3-dimensional z-score maps of brain parenchymal TSPO-PET were generated by voxel-by-voxel statistical comparison between each subject’s normalized PET images and a HC dataset. Given that the initial SUV-based evaluation showed a global decrease in [F-18]PBR06 uptake in the brain following nasal foralumab at three months in the index case, global normalization was not considered to be an appropriate approach to detect treatment effects of nasal foralumab. For initial alternate analysis and as a proof of concept, normalization was performed using a white matter pseudoreference region based on PET voxels showing the lowest quartile of 60-90-minute SUVs in the index-case (GALP 2.0 or p-GALP approach) and the z-score images hence generated were used for initial visual interpretation. Subsequently, for additional unbiased analyses, normalization was performed against a modified pseudoreference region developed in-house by our group, based on white matter voxels showing no significant differences between MS patients and healthy controls in our historical cross-sectional cohort, similar to approaches adopted by other investigators for other PET tracers (GALP 3.0 or m-GALP approach).^30, 31^ Global, cortical, white matter, thalamic and cerebellar regions of interest (ROIs) were defined in the atlas space and average z-scores from images generated using the above approaches (referred to as GALP2.0 (or p-GALP) and GALP 3.0 (or m-GALP) approach, were extracted in these ROIs using PMOD.^29^ Global ROI represented a composite region comprising of the cortical, white matter, thalamic and cerebellar ROIs.

### 3T MRI Acquisition and Analysis

All subjects underwent brain and spinal cord MRI scans on the same scanner (Siemens 3T Skyra, Siemens Healthineers, Erlangen, Germany) using the same acquisition protocol. Whole-brain images were acquired at high-resolution (voxel sizes 1mm3) with 3D magnetization-prepared rapid gradient-echo (MPRAGE), 3D T2-weighted, and 3D T2 fluid-attenuated inversion recovery (FLAIR) sequences as previously reported.^32^ Cervical and thoracic spinal imaging was obtained with sagittal T1 spin-echo, T2 spin-echo, short tau inversion recovery (STIR), and post-contrast sagittal and axial T1 spin-echo sequences; full sequence details have been previously published.^33^ Within 6 months prior to the start of foralumab, patients underwent MRI of the brain, cervical and thoracic spine with administration of intravenous gadolinium contrast to confirm the absence of new/enlarging T2 or gadolinium-enhancing lesions prior to enrollment. Brain MRI was performed at approximately 3 months after treatment start, and complete brain, cervical and thoracic spine MRI was obtained at approximately six months. For these three-and six-month follow-up scans, gadolinium was not administered. An expert reader (JZ, TC) analyzed the MRI scans for the presence of new or enlarging T2 lesions.

### Lumbar puncture

A lumbar puncture for clinical purposes was conducted in EA3-11 at baseline and 6 months. Cerebrospinal fluid was evaluated for cell count, oligoclonal bands and viral testing.

### Statistical Methods

To estimate the change after treatment in the EDSS, MSFC-4, MFIS, and NeuroQoL, the change over the subsequent six- and twelve-month periods, were calculated for each outcome for subjects with complete data, and a Wilcoxon signed rank test was used to calculate the p-value for the null hypothesis of no change over time. When proportions were calculated, 95% confidence intervals were based on the exact binomial distribution. All statistical analysis was completed in the statistical package R version 4.3.0 (www.r-project.org).

### Proteomic analyses

Olink analysis: A 48-Plex by Olink^®^ using Proximity Extension Assay (PEA) technology was conducted in the serum and CSF.^29^ We compared changes in proteins across different timepoints using a linear mixed effect model (random intercepts) adjusting for plate using the nlme library in R. All comparisons were adjusted for multiple testing using the holm’s method. SIMOA analysis: sNFL and GFAP by Quanterix^™^ using the SIMOA assay as previously described.^34^

### Single cell RNA sequencing

<u>RNA sequencing</u>: Peripheral blood mononuclear cells (PBMCs) from EA3-6 at baseline, three, and six months were processed for scRNAseq. PBMCs were thawed and processed into single cell suspensions using established methods. Initial cell viability measurements from all vials were greater than 90%. Cell suspensions were loaded onto a Chromium Single Cell 3’ Chip (10x Genomics, Inc.) and processed into Illumina-compatible libraries using the Chromium Single Cell 3’ dual index (v3.1 Chemistry) kit according to the manufacturer’s protocol. Sequencing was performed on a NovaSeq 6000 platform. Raw data was processed using cellranger v6.0 and input into R/Seurat (v5.0.1) for downstream analysis.

<u>Whole genome sequencing and Demultiplexing</u>: DNA corresponding to PBMCs from each of the four individual patients was extracted using the Qiagen DNeasy Blood & Tissue Kit. Extracted gDNA was converted into sequencing libraries using the NEBNext Ultra II FS DNA Library Prep Kit. Libraries were sequenced on a NovaSeq 6000 platform. Germline variants were called from raw sequencing data using the Sarek workflow following GATK best practice guidelines. A “cohort” GVCF file was generated for use in demultiplexing the pooled scRNA-seq data.

<u>scRNAseq data analysis</u>: Using Seurat,^35–37^ genes were filtered for only those which were present in three or more cells, and cells were filtered for those which had at least 200 genes and <10% of reads mapping to mitochondrial genes. Data were then normalized and scaled, before integration using the Harmony method.^38^ Genetic demultiplexing was performed using souporcell;^39^ any cells not mapping to a subject or which mapped to multiple subjects were removed. Cell type assignments were called using Azimuth utilizing the PBMC reference^40^ provided by the Satija lab.

Differential expression analysis was, limited to genes which have 100 reads or more. Using AggregateExpression and FindMarkers functions with the DESeq2 method,^41^ comparisons were run of overall expression, and expression by cell type comparing the three- and six-month time points back to the baseline expression levels. These the DEG lists were then used to perform GSEA using the MSigDB Hallmark gene set^42^ and fgsea (https://doi.org/10.1101/060012) in R.

## RESULTS

### Baseline characteristics of patients

Baseline characteristics are shown in **Table 1**. There was one screen failure due to nasal exam findings that precluded sufficient drug delivery.

**Table 1:** Baseline characteristics of expanded access patients, and EDSS after 6 and 12 months of continuous dosing.

| Subject | Sex | Age at treatment start | Race | Ethnicity | Last MS Treatment | Treatment Start | Dosing cycles completed | Baseline EDSS | 6-month EDSS | 6 month Change | 12 month EDSS | 12 month change |
| --- | --- | --- | --- | --- | --- | --- | --- | --- | --- | --- | --- | --- |
| 1 | M | 60-65 | White | Non-HL | Ocrevus 12/2020 | 05/2021 | 21 | 6.0 | 6.0 | 0 |  |  |
| 2 | M | 40-45 | White | Non-HL | Ocrevus 10/2021 | 01/2022 | 38 | 6.0 | 6.0 | 0 | 5.5 | -0.5 |
| 3 | F | 65-70 | White | Non-HL | Ocrevus 04/2022 | 01/2023 | 28 | 6.5 | 6.5 | 0 |  |  |
| 4 | M | 50-55 | White | Non-HL | Ocrevus 12/2020 | 12/2022 | 23 | 2.5 | 2.0 | -0.5 |  |  |
| 5 | F | 60-65 | White | Non-HL | Ocrevus 01/2022 | 01/2023 | 28 | 6.0 | 6.0 | 0 |  |  |
| 6 | F | 55-60 | U/NR | NR | Ocrevus 11/2021 | 01/2023 | 8 | 6.0 | 6.0 | 0 |  |  |
| 7 | F | 60-65 | White | Non-HL | Ocrevus 06/2023 | 09/2023 | 22 | 6.5 | 6.5 | 0 | 6.5 | 0 |
| 8 | F | 60-65 | White | Non-HL | Rituxan 02/2023 | 08/2023 | 23 | 3.5 | 3.0 | -0.5 | 2.5 | -1.0 |
| 9 | M | 60-65 | White | Non-HL | No DMT | 09/2023 | 18 | 4.5 | 3.5 | -1.0 | 3.5 | -1.0 |
| 10 | M | 40-45 | Black | Non-HL | Ocrevus 03/2023 | 10/2023 | 13 | 6.5 | 6.5 | 0 |  |  |

Legend: HL – Hispanic/latino; NR=not reported

### Treatment periods

(**Supplementary** Figure 1) Patient 1 was treated over 1.8 years with breaks, then discontinued due to an unrelated medical issue that made him ineligible. Patients 2, 7, 8, and 9 were treated continuously for over a year and are ongoing. Patients 3-6 were treated for six months, after which patient 3, 4, 5 had a treatment hiatus for approximately three months, then resumed and are ongoing. EA6 discontinued after six months due to pseudorelapses without MRI evidence of new lesions and environmental allergies. Patient 10 was treated for nine months, then discontinued after a fall at month eight resulting in a quadriceps tear.

### Adverse events (AEs)

There were 206 treatment emergent adverse events (TEAEs) (**Supplementary Table 4**) of which 34 were treatment related (TRAEs) (**Supplementary Table 5**). Adverse events are reported by the patient or through an exam or laboratory finding. There was one episode of tingling and numbness in the lips and tongue which was deemed related to treatment, and this patient was treated with cetirizine prior to dosing which alleviated this symptom. After 11 cycles, cetirizine-pre-treatment was suspended, and the patient continued treatment without incident. Two patients discontinued treatment, one due to an orthopedic tear which was unrelated to the drug, and the other due to pseudorelapses and environmental allergies, which were unrelated to treatment. Both resumed B-cell therapy. No treatment-related SAEs were reported. Two patients were hospitalized for COVID in 2022; both had uneventful recoveries There were no deaths.

### 3T MRI

Standardized non-contrast 3T MRIs of the brain and spine on the same Siemens scanner were conducted at baseline and every three months. None of the patients demonstrated any new T2 lesions over the treatment period.

### EDSS

Since all patients completed six months of continuous dosing, we first evaluated EDSS changes at the six-month timepoint (**Table 1**). All patients stabilized, and three out of 10 improved in their EDSS scores. One of 10 improved using a more stringent definition of EDSS improvement (EDSS≤5.5, improvement of 1.0; EDSS >5.5, improvement of 0.5). In four patients continuously treated for 12-months, three had improvement in EDSS, and the fourth had no change (**Table 1**).

<u>MSFC4</u>: MSFC4 was conducted every three weeks. At six months, there was no overall change in the Timed 25-Foot Walk (T25FW) or Nine Hole Peg Test (9-HPT) in the dominant and non-dominant hand. The SDMT improved by≥3 points in nine out of 10 patients, likely due to practice effect. Interestingly, there was significant improvement (p = 0.049) in binocular LCLA, with four out of 9 patients having an improvement of ≥8 points. (**Supplementary Table 1**).

### MFIS and NeuroQoL

MFIS questionnaires were conducted every three weeks. We report the mean change from baseline to 6 months in the MFIS total score and subscores in **Table 4A**, demonstrating trends in score reductions. A four-point change in MFIS has been determined to be clinically significant.^43^ Six out of 10 patients demonstrated a reduction in total MFIS of four points or more over the six-month period. Of the four patients with continuous dosing over a 12-month period, 100% experienced a >4 point reduction in total MFIS at 12 months compared to baseline (**Table 4B**). A short

NeuroQoL questionnaire was administered every three months. There was no significant change in the subscores from baseline to six months (**Supplementary Table 2**).

### TSPO-PET

Widespread increased radiotracer uptake was observed in the brain of the index case (EA1) on evaluation of the baseline standardized uptake value (SUV) image. Following treatment, there was a marked reduction in [F-18]PBR06 uptake throughout the brain at three months (**Supplementary** Figure 3). Similar reductions were seen in other subjects (**Supplementary** Figure 3A). Individualized evaluation of abnormal z-score maps revealed widespread abnormal voxels in subjects prior to treatment that decreased with treatment. (**Figure 1A, B, Supplementary** Figure 4B).

**Figure 1A and 1B.**
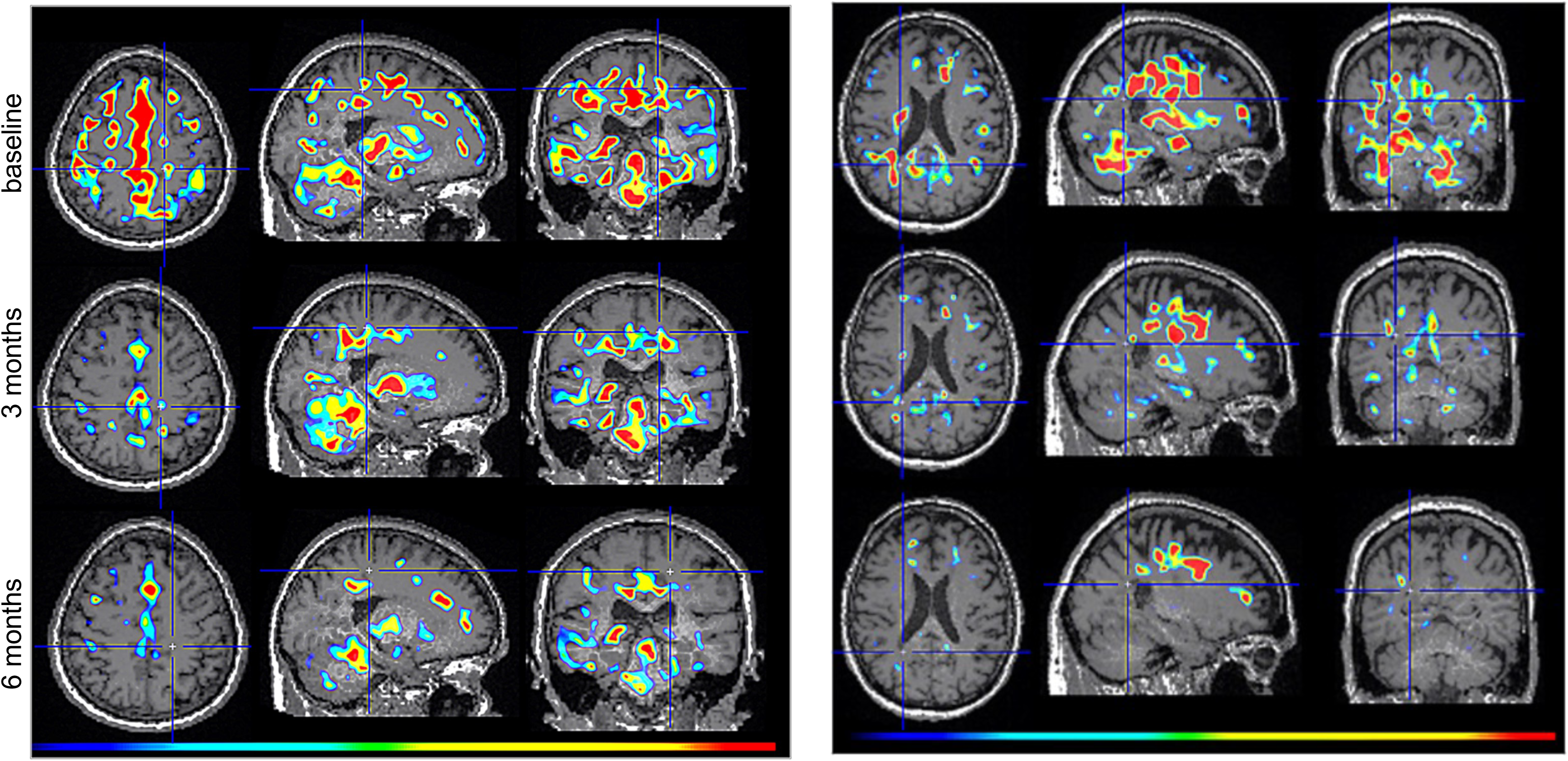
Reduced TSPO-PET signal following nasal foralumab treatment. Individualized pGALP z-score maps for subjects EA6 (**A**) and EA1 (**B**) superimposed on respective subject’s T1-weighted MRI demonstrate decreased PET abnormalities following treatment at 3 months (*middle row*) and 6 months (*bottom row*) as compared to baseline (*top row*). Hotter colors represent a higher z-score value and cooler colors represent a lower z-score value as determined by the GALP 2.0 pipeline. GALP=Glial activity load on PET.

**Figure 1C and 1D.**
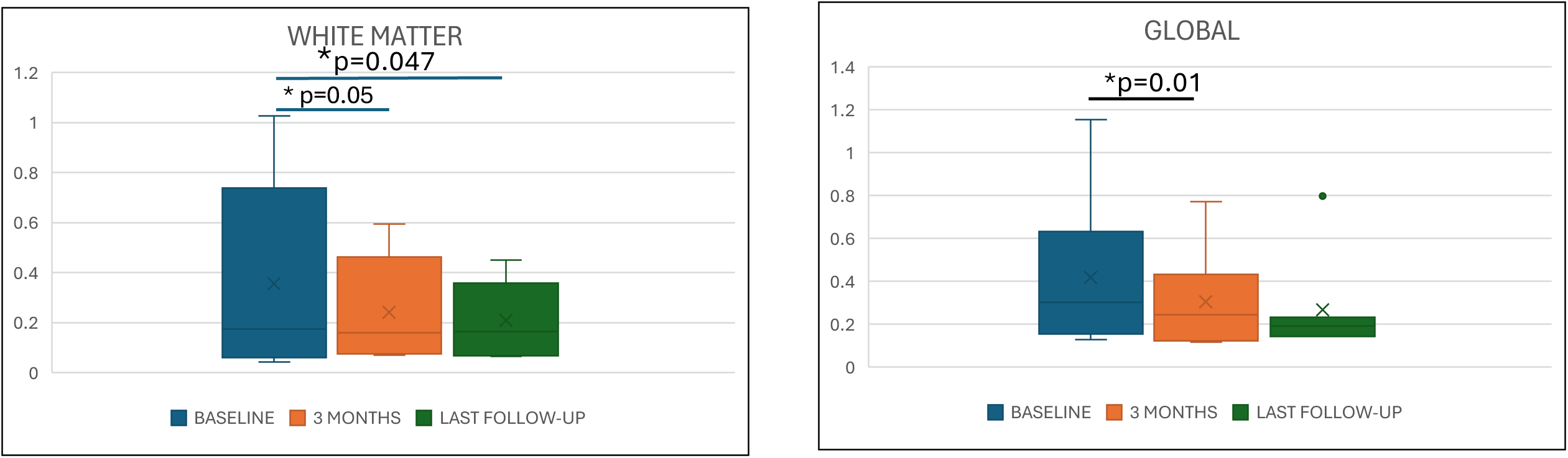
Decreased voxel-wise average [F-18]PBR06-PET mGALP z-scores (GALP 3.0) in white matter (**C**) and global (**D**) regions of interest following nasal foralumab treatment for 3 months and 7.5 months (mean duration of treatment at last available PET follow-up visit).

**Figure 1E and 1F.**
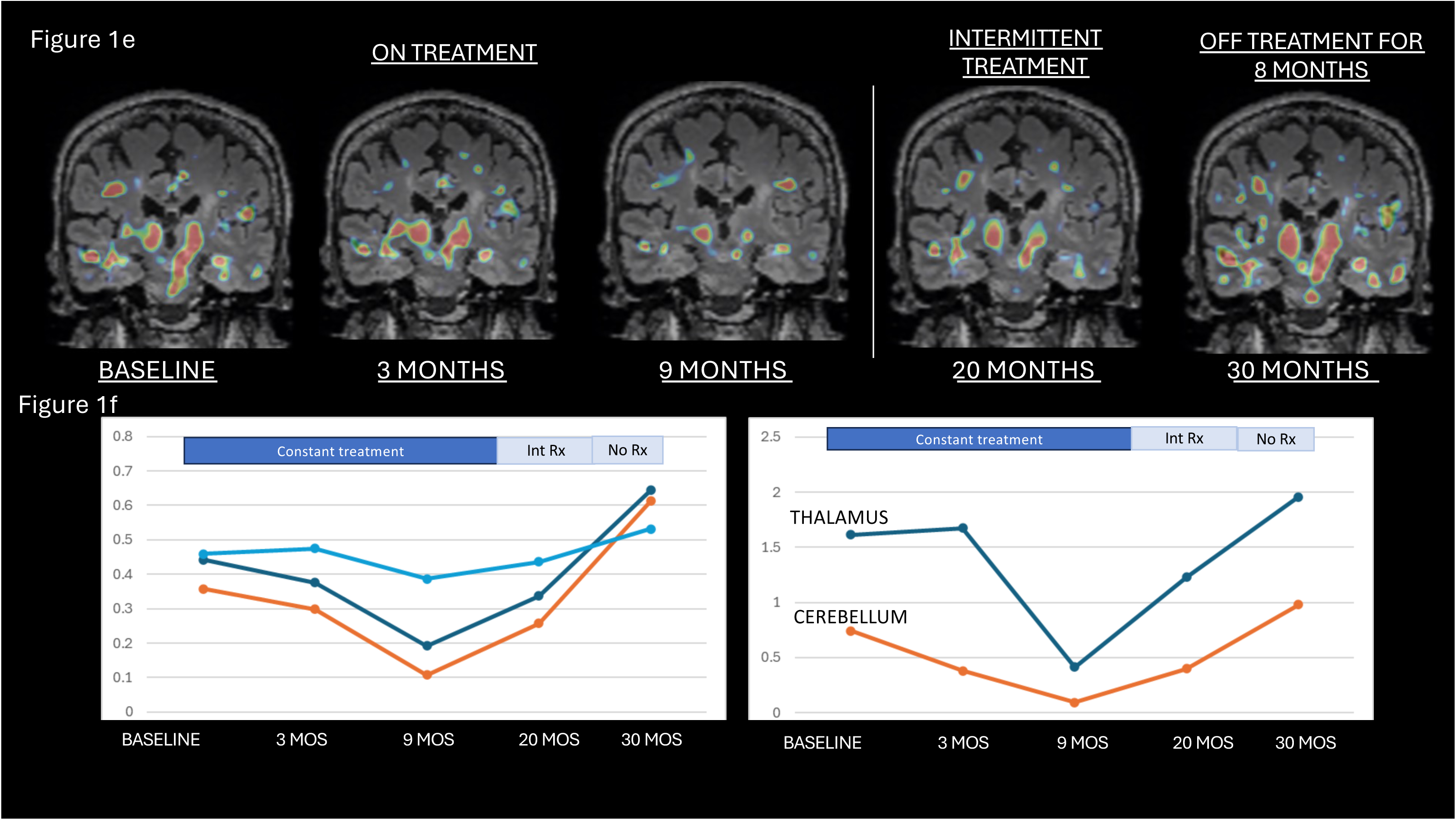
Individualized mGALP z-score (GALP 3.0) maps for EA1 superimposed on subject’s T2-FLAIR MRI demonstrated decreased glial activity load on PET while the patient was on treatment with nasal foralumab. PET scans performed after patient was on intermittent treatment and subsequently went off treatment showed a progressive increase in glial activity (**E**), confirmed by quantitative evaluation across brain regions (**F**).

Quantitative group analysis after three months of treatment demonstrated a reduction in average PET mGALP score in the white matter ROI (0.40 +/-0.09 versus 0.47 +/-0.09, −14.9%, effect size 0.875, p=0.055) and the global ROI (0.31+/-0.23 versus 0.42 +/-0.36, −26.2%, effect size 0.78, p=0.016). (**Table 3**. **Figure 1C, D**). Quantitative analysis at the time of the last PET scan on treatment (average duration 7.5 months) demonstrated a reduction in white matter (0.38+/-0.09 versus 0.47+/-0.09, −19.1%, effect size 0.89, p=0.047, respectively, n=7) and global (0.26+/-0.24 versus 0.42+/-0.36, −38.1%, effect size 0.73, p=0.11, n=7) ROIs (**Table 3**, **Figure 1C, D**). Six-month change as compared to baseline is depicted in **Supplementary Table 3**.

**Table 2:** Total Modified Fatigue Impact Scale (MFIS) and subscores in 10 subjects over 6- and 12-month continuous foralumab dosing periods.

| 6 month values | Baseline<br>(mean, SD) | 6 month<br>(mean, SD) | Change | Improvement by >4<br>points on total MFIS |
| --- | --- | --- | --- | --- |
| **MFIS total** | 30.0 (10.1) | 24.1 (15.5) | -5.9 (11.3) p=0.16 | 6/10 |
| **MFIS Cognitive** | 6.1 (5.6) | 4.3 (5.0) | -1.8 (5.1) p=0.24 |  |
| **MFIS Physical** | 20.3 (6.6) | 16.8 (9.1) | -3.5 (6.8) p=0.19 |  |
| **MFIS Psychosocial** | 3.6 (1.8) | 3.0 (2.9) | -0.6 (2.0) p=0.43 |  |
| 12 month values | Baseline<br>(mean, SD) | 12 month<br>(mean, SD) | Change | Improvement by >4<br>points on total MFIS |
| **MFIS total** | 26.8 (11.0) | 12.3 (9.3) | -14.5 (6.6) p=0.13 | 4/4 |
| **MFIS Cognitive** | 3.0 (2.2) | 0.0 (0.0) | -3.0 (2.2) p=0.13 |  |
| **MFIS Physical** | 20.8 (9.7) | 11.0 (7.4) | -9.8 (5.3) p=0.13 |  |
| **MFIS Psychosocial** | 3.0 (2.6) | 1.3 (1.9) | -1.8 (2.1) p=0.20 |  |

**Table 3:** mGALP scores for baseline, 3 months and the last PET scan (average duration 7.5 months) on treatment.

| A | Baseline | 3-month | Change | Wilcoxon signed rank p-value |
| --- | --- | --- | --- | --- |
| Global | 0.42 (0.36) | 0.31 (0.23) | -0.11 (0.14) | 0.016 |
| Cortex | 0.37 (0.47) | 0.25 (0.28) | -0.12 (0.20) | 0.11 |
| Thalamus | 0.99 (0.90) | 0.84 (0.61) | -0.15 (0.44) | 0.46 |
| White matter | 0.47 (0.09) | 0.40 (0.09) | -0.07 (0.08) | 0.055 |
| Cerebellum | 0.49 (0.60) | 0.33 (0.57) | -0.16 (0.20) | 0.11 |
| B | Baseline | Last visit* | Change* | Wilcoxon signed rank p-value* |
| Global | 0.42 (0.36) | 0.26 (0.24) | -0.11 (0.15) | 0.11 |
| Cortex | 0.37 (0.47) | 0.19 (0.28) | -0.12 (0.21) | 0.30 |
| Thalamus | 0.99 (0.90) | 0.81 (0.66) | -0.14 (0.87) | 0.69 |
| White matter | 0.47 (0.09) | 0.38 (0.09) | -0.067 (0.075) | 0.047 |
| Cerebellum | 0.49 (0.60) | 0.25 (0.55) | -0.20 (0.26) | 0.16 |

*Legend: \*One subject was removed from the last-visit point and last-visit change analysis*

Nasal foralumab was associated with a major effect on reduction of fatigue in most patients treated (**Table 2A, B**). Thus, we analyzed the relationship between MFIS scores and microglial PET signal. We focused on the hippocampus and the substantia nigra, as functional and neuroinflammatory changes in these regions have been associated with fatigue in MS.^44^ Total MFIS scores correlated strongly with mGALP scores in the hippocampus (r=0.89, p=0.007) at baseline (**Supplementary** Figure 5). Among the subjects with total MFIS>30 or cognitive MFIS>10 (n=5), there was a significant reduction in PET mGALP scores in the hippocampus and substantia nigra following nasal foralumab treatment (p<0.05) (**Supplementary** Figure 6).

After observing a decrease in the global and white matter PET signal after treatment, we asked whether the PET signal specifically associated with perilesional white matter lesions also changed with treatment. As shown in **Supplementary** Figure 7 there was a clear reduction in perilesional PET signal following treatment seen at three and six months in individual subjects.

Patient 1 (EA1), the first subject treated, had the longest microglia PET follow up, including scans performed at baseline and at three, nine, 20 and 30 months from the time of treatment initiation. At 20 months he received intermittent treatment and at 30 months he was off treatment for eight months. This allowed us to investigate the microglial PET signal while on treatment, while receiving only intermittent treatment and when he was off treatment for eight months. As shown in **Figure 1E and 1F**, there was a reduction of PET signal while on treatment with an increase back to baseline after stopping treatment.

### Blood biomarkers

Cytokines and inflammatory markers in the serum were assayed using the Olink proximity extension assay platform (48-plex) at baseline (T1), three months (T3) and six months (T6). We found that foralumab treatment resulted in a reduction in IL-7, CCL2, FLT3lg and VEGFa (p<0.05) over the six-month period (**Figure 2**). We measured serum NfL and GFAP SIMOA assay over the same three timepoints and found no change in these biomarkers.

**Figure 2.**
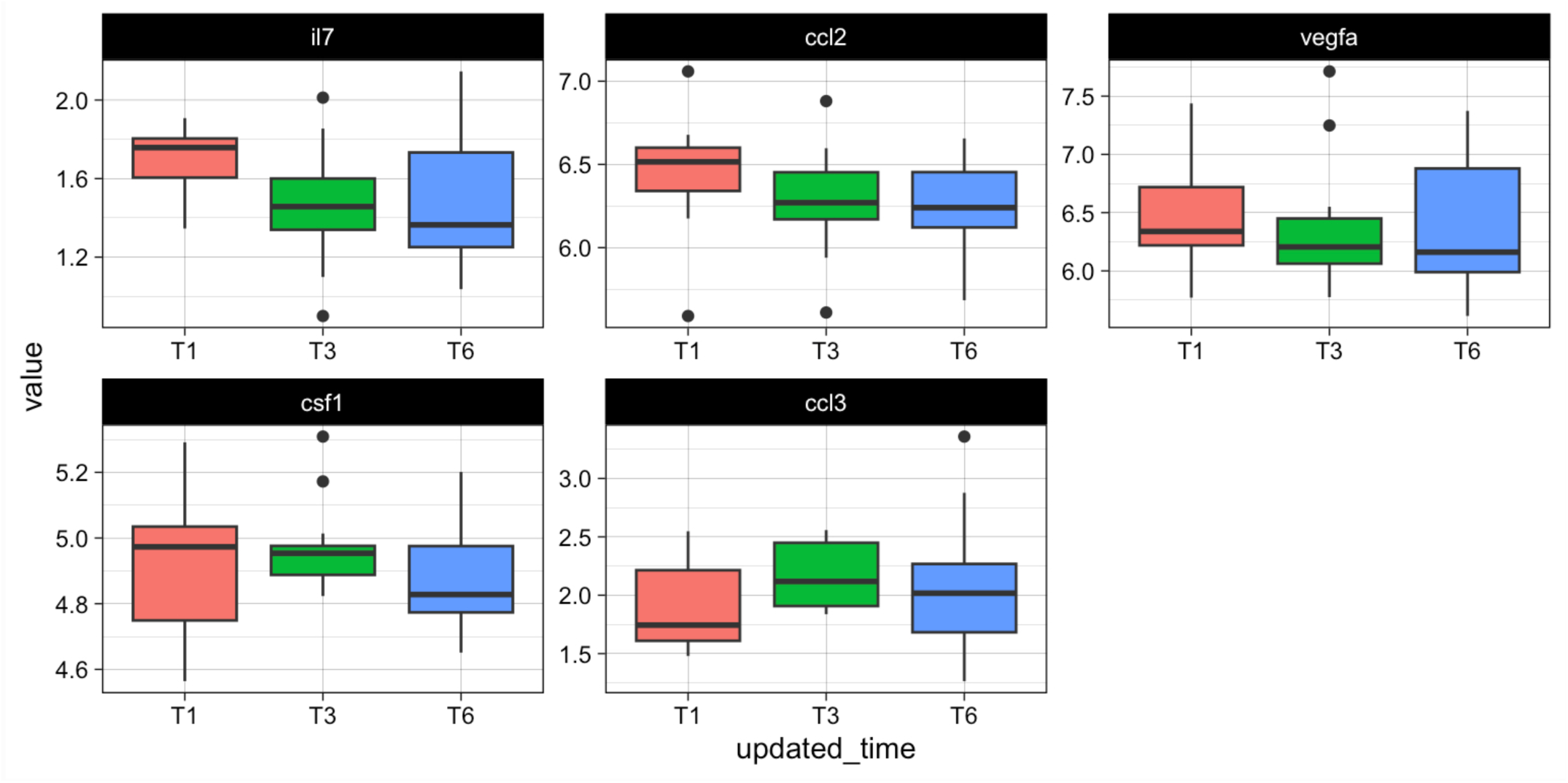
Olink 48-plex cytokine analysis in the serum of foralumab-treated patients at baseline (T1), 3 months (T3) and 6 months (T6), shows a reduction in IL-7, CCL2, FLT3lg and VEGFa *p<0.05.

Anti-drug antibodies: Anti-drug antibodies (ADAs) were evaluated in serum samples from EA1 and EA2 (baseline, three months, four-five months, nine months, 12 months and 15-17 months, as well as EA3-6 baseline, three, and six months), and no ADAs were found in any samples.

### CSF

Lumbar punctures were conducted at baseline and six months in six patients. Two patients had only one LP due to access issues. Total nucleated cell number ranged between 0-6 for all LPs. There was no significant change in the number of oligoclonal bands at 6 months compared to baseline (**Supplementary** Figure 1). Olink 48-plex cytokine analysis in the CSF of foralumab-treated patients at baseline (T1) and six months (T2), showed a trend in reduction of CXCL12 (**Supplementary** Figure 2B).

<u>scRNAseq:</u> scRNAseq of PBMC samples at baseline, three and six months in four subjects shows changes in overall gene expression at the three-month timepoint and sustained at six months (**Figure 3**). Gene expression was significantly altered in the following cell subsets: NK cells, CD14+ monocytes, CD16+ monocytes, CD4+ central memory cells (Tcm), regulatory T cells (Tregs), naïve B cells, CD8 effector memory cells (Tem), conventional dendritic cells (cDCs) and central memory CD8 cells (Tcm) as shown by heatmap of differential gene expression in cell subsets with the most significant changes at three and six months compared to baseline (**Figure 4**). Genes which were differential at six months also contributed to pathway differences, which varied by cell type, but included a robust decrease in interferon-alpha pathways (**Figure 5**). We identified three key differentially expressed genes in these six cell subtypes, representing critical pathways involved in MS pathogenesis: Beta-2 microglobulin (B2M) which associates with MHC class I molecules was decreased at three and six months in all six cell types (**Figure 6**); STAT1 was reduced in naïve B cells, CD4 Tcm and CD8 Tem, and Foxp3+ Tregs at three months, and Tregs at six months compared to baseline; TGF-beta 1 (TGFB1) was increased in CD14+ monocytes, CD16+ monocytes, naïve B cells, CD4 Tcm and CD8 Tem.

**Figure 3.**
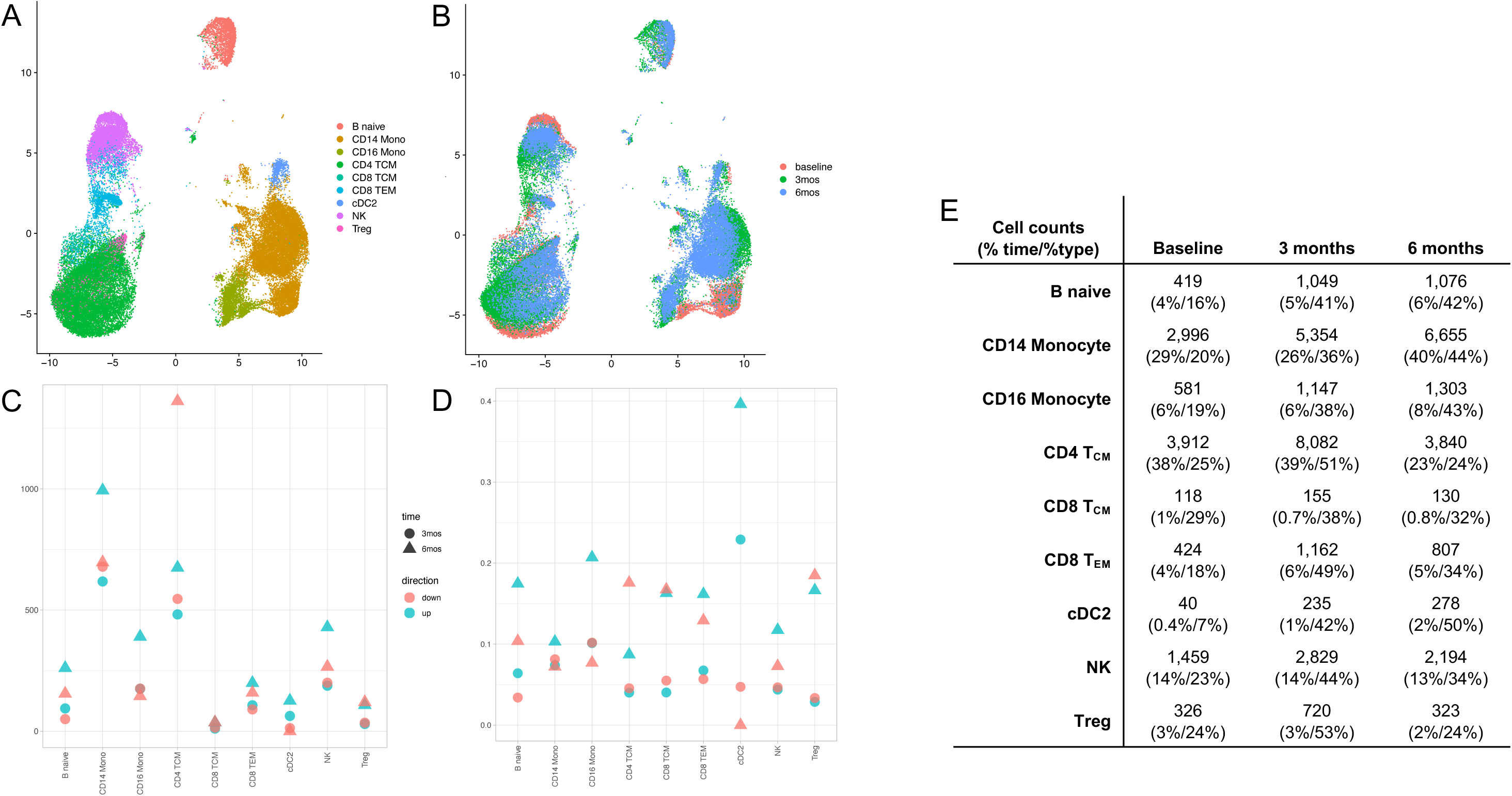
A. scRNA UMAP of cells labeled by celltype. **B** same cells as A, but color indicates sample time point. Samples were sequenced in batches corresponding with sample time point but were fully processed in tandem. **C** Raw counts of DEGs by cell type, shape indicates time point, and color indicates log2FC direction, versus baseline. **D** normalized counts of DEGs by cell type, shape indicates time point, and color indicates log2FC direction, versus baseline. **E** Table of counts of cells by type and time point.

**Figure 4.**
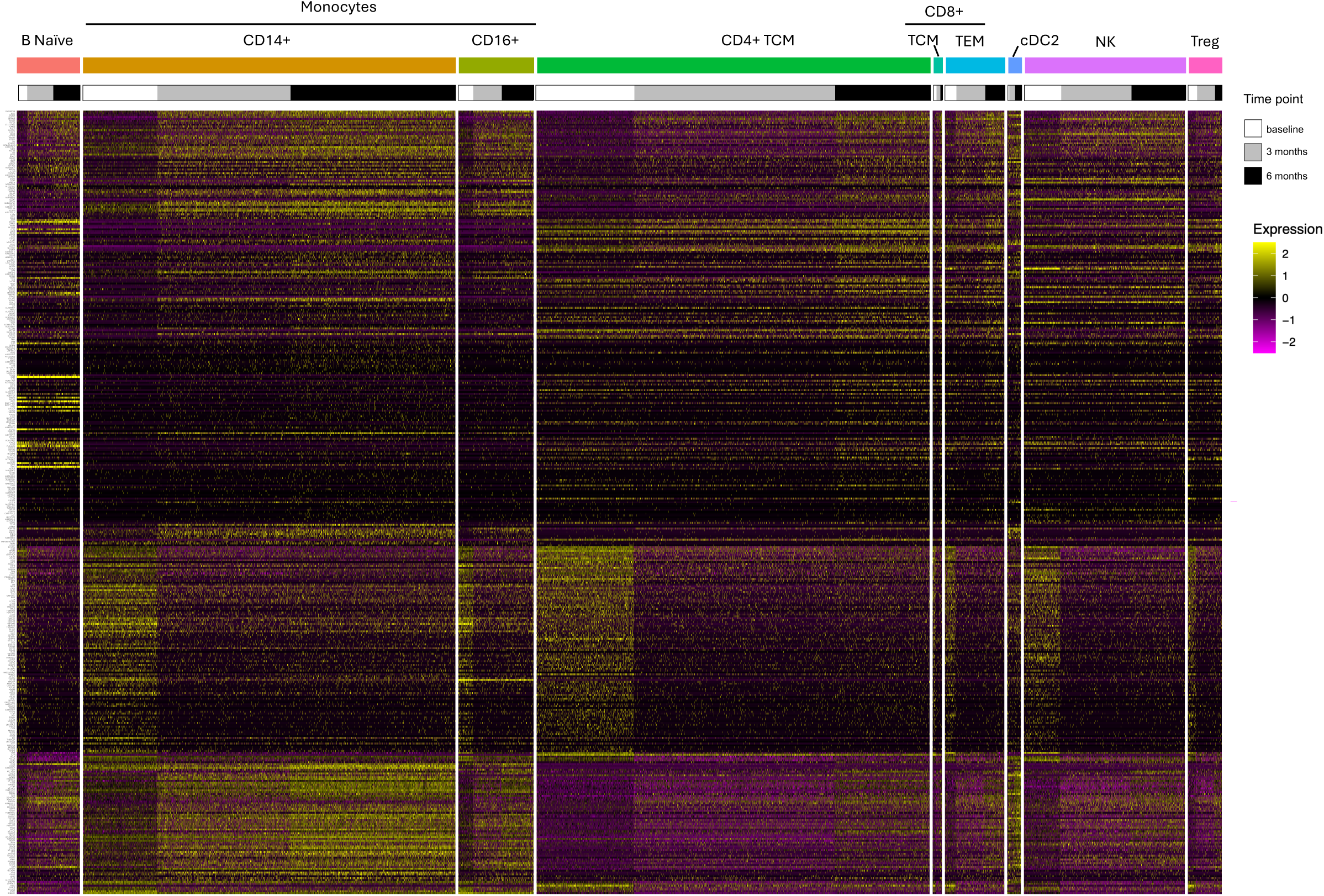
Heatmap with cells as columns, including only those genes which have |average log2FC| > 2 and a Bonferroni adjusted p-value < 0.001 in at least one cell type, total of 552 genes plotted. Colors across the top indicate cell type, colors across the bottom indicate timepoints. Expression values are scaled.

**Figure 5.**
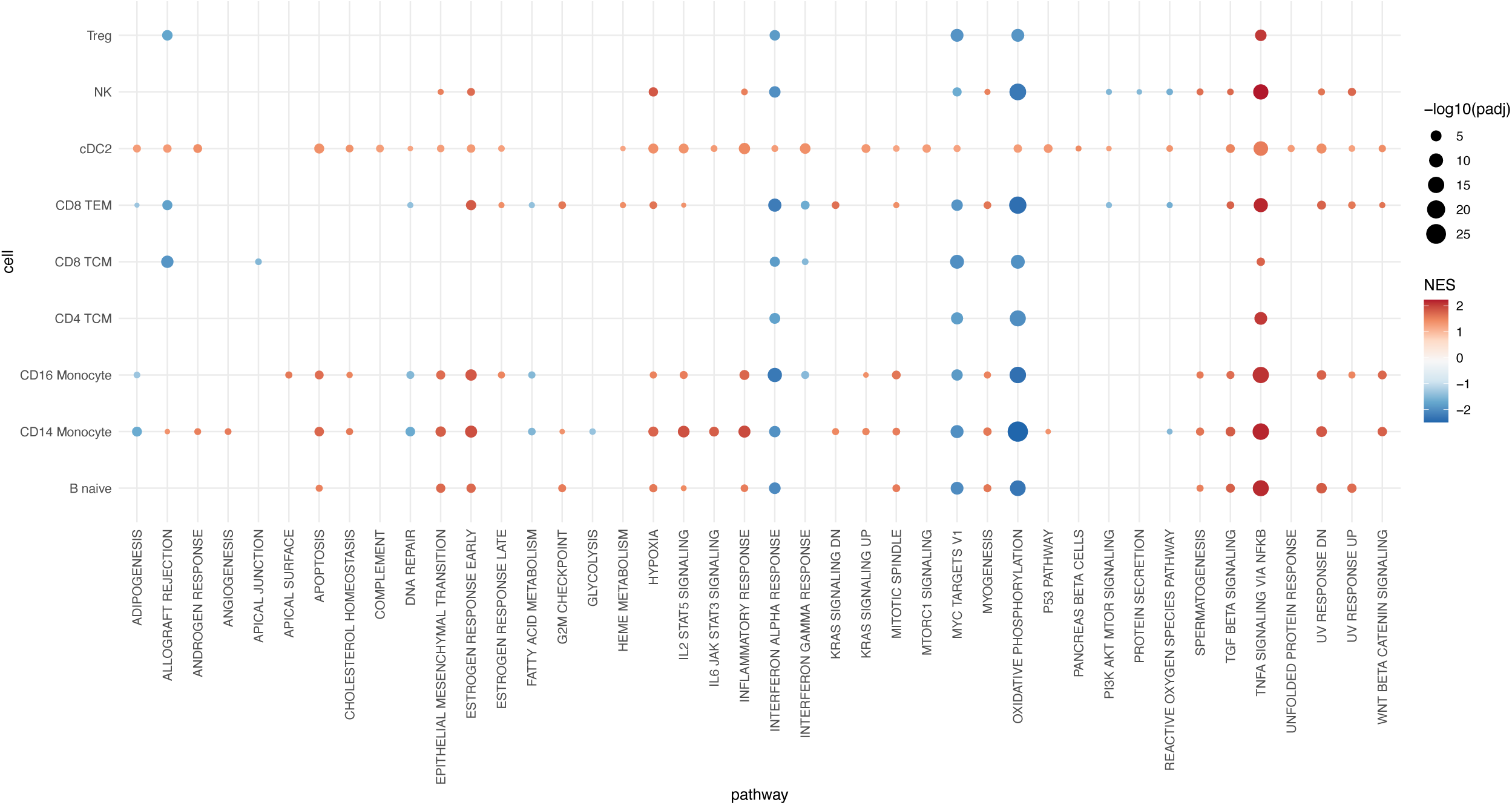
Pathway enrichment was run using fgsea, using the DEGs from the 6 months versus baseline comparison for each cell type. Only those pathways which were significant in at least one cell type are included. Color indicates normalized enrichment score (NES), and size is −log10(adjusted p-value) of the enrichment.

**Figure 6.**
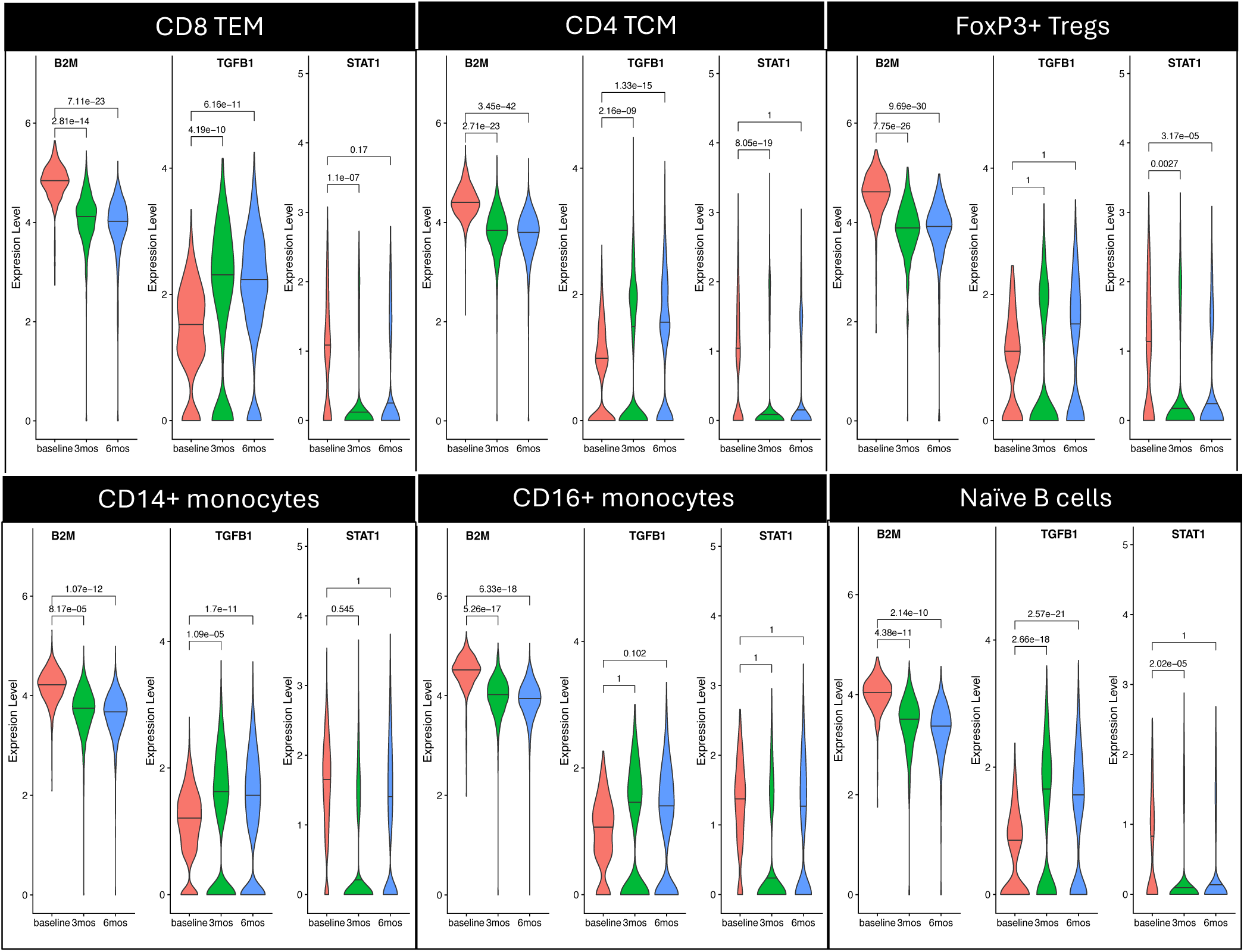
Violins include expression from all cells at given time point in given cell type. Horizontal line through each violin indicates the median expression value. Included values on significance bars are the FDR adjusted p-values of the differential expression.

## DISCUSSION

We treated 10 naSPMS patients with nasal foralumab in this expanded access program for at least six months, and one for 1.8 years. Clinically, we found that all patients stabilized in terms of their EDSS scores, and three out of four patients treated for 12 months continuously demonstrated improvement on their EDSS. Six out of ten patients demonstrated an improvement in fatigue scoring on the MFIS scale. There were no treatment-related SAEs or severe AEs. There was a significant reduction in WM-PET TSPO signal over six months, as well as a significant increase in TGF-beta expressing T cells in circulation, consistent with effects seen in our EAE study of anti-CD3.^14^ These findings support further studies to evaluate the effects of nasal anti-CD3 in progressive forms of MS and in patients with PIRA.

Nine of the 10 patients were previously treated with B cell depleting therapy and were worsening despite therapy. After starting nasal foralumab, the EDSS score stabilized in all patients, and none worsened over the six-month period, with some patients experiencing an improvement in EDSS after longer-term treatment. Although these results are promising, this is an unblinded open-label study, and clinical efficacy requires evaluation in placebo-controlled studies. A phase 2a randomized double-blind placebo-controlled study is in progress (NCT06292923).

A four-point reduction in the total MFIS score is clinically significant.^43^ Fatigue measured by the MFIS score improved by at least four points in six out of 10 patients at six months. We have found fatigue to correlate with PIRA (Molazadeh, in preparation) suggesting similar biological mechanisms. Our other studies have found fatigue to be correlated with the TSPO signal in the multiple brain regions including substantia nigra^23^ and our recent analyses suggest that increased microglial activation may mediate the relationship between fatigue and PIRA,^45^ and PIRA may be mediated by similar mechanisms, including CNS-centric inflammation. In the current study, we saw a significant reduction in fatigue scores following nasal foralumab treatment among the subjects who had baseline fatigue, which was accompanied by a reduction in TSPO-PET signal in substantia nigra and hippocampus. This finding has several important implications: 1) nasal foralumab may have a clinically significant impact on fatigue, a highly disabling symptom, in naSPMS patients; 2) the effect of nasal foralumab on fatigue may be mediated by its effects on microglial activation; and 3) this finding provides further longitudinal evidence for a causal relationship between microglial activation and fatigue in MS.

While multiple, cross-sectional studies have shown increased TSPO-PET uptake in progressive as compared to relapsing MS, no PET studies have studied treatment effects in an exclusive population of naSPMS subjects. One study evaluated the effects of natalizumab on a mixed population of relapsing remitting and secondary progressive MS and showed that there was less than 5%, albeit statistically significant reduction, in distribution volume ratio of [C-11]PK11195-PET in the treated population, as compared to an increase in the untreated population.^46^ In other treatment-related PET studies in RRMS, fingolimod and glatiramer acetate resulted in a 3-12% reduction in [C-11]PK11195-PET.^47, 48^ While the various PET measurement indices across tracers, patient populations, multiple centers and differing methodologies may not be comparable, we believe that 15-38% TSPO-PET reduction in our study of treatment-refractory, naSPMS patients following nasal foralumab represents a significant treatment effect. We have also found marked reduction of TSPO-PET signal in an Alzheimer’s patient treated with nasal foralumab.^49^

These studies provide unique human data supporting our group’s preclinical findings where we have shown that nasal anti-CD3 decreased microglial activation in animal models of neurologic diseases including progressive MS,^14^ Alzheimer’s disease^50^ and traumatic brain injury.^51^ Of note, nasal anti-CD3 affects non-CNS diseases as well including lupus^10^ and diabetes.^52^

Our longitudinal PET data in the index subject (EA1) collected over 30 months showing a progressive increase in PET signal after the subject stopped nasal foralumab, serves as a within-subject control, and provides additional evidence that the reduction in PET signal during treatment was likely related to nasal foralumab’s effects.

Overall, the treatment was well-tolerated and there were no treatment-related SAEs or severe adverse events. Consistent with the tolerability of antibodies given by the mucosal route, no side effects were observed in colitis subjects given oral anti-CD3 (OKT3).^53^ Of note, intravenous anti-CD3 has been associated with EBV activation and anti-drug antibodies.^12^ The majority of TRAEs were nasal or respiratory in nature, and included nasal congestion, runny nose and abnormalities on nasal endoscopy examination. None of the patients discontinued treatment due to nasal symptoms or abnormalities. One patient failed screening due to nasal anatomy that would limit optimal drug delivery. Thus, not all patients may be eligible for a long-term nasal therapy, or other methods of nasal delivery may need to be developed. There was no increase in infections in nasal foralumab treated patients, some treated as long as a year. In animal studies, nasal anti-CD3 treatment did not interfere with ability of the lung to clear a bacterial infection^14^ or interfere with clearance of lung infection in COVID patients treated with nasal foralumab.^54^

Nasal foralumab resulted in changes in key gene expression by single cell RNA sequencing in monocytes and CD8 T cells including a reduction in beta-2 microglobulin (B2M) and an increase in the regulatory molecule TGFβ1 as early as three months post treatment in a variety of cell types including T cells and monocytes, leading to an immunoregulatory, tolerizing environment. These results are consistent with our prior studies demonstrating an increase in TGFβ1 in healthy controls and patients with COVID-19 treated with nasal foralumab^54^ as well as our murine studies with nasal anti-CD3 which resulted in TGFβ1-expressing regulatory T cells within the CNS and suppression of disease in a progressive model of MS.^14^ Thus, the induction of TGFβ1 by nasal foralumab in multiple of cell types may be a key mechanism by which it suppresses innate inflammation in the CNS of MS patients as was shown in the murine model. Studies of teplizumab (anti-CD3) given intravenously for type 1 diabetes showed a correlation of change in frequency of exhausted CD8+ T cells (CD8+KLRG1+TIGIT+) with the fold change in C-peptide at month 6, as well as a reduction in IFNγ expression.^55^ We observed a reduction in STAT1 in CD4 and CD8 T cell subsets in response to nasal foralumab accompanied by a reduction in type I and type II interferons. In terms of serum proteomic biomarkers, we found a reduction in CCL2 (MCP-1) which recruits monocytes, dendritic cells and T cells to the sites of inflammation,^56^ consistent with the reduction in microglial TSPO-PET signal in the CNS of treated patients.

Limitations: Because this was an open label study, no conclusions can be made about clinical effects, which will require a placebo-controlled double blind, which is ongoing. Our current microglia PET measure is a newly developed measure and use of this approach in the evaluation of microglial PET findings will require further evaluation in other subjects incorporating additional quantitative and semi-quantitative approaches.

Treating naSPMS is an unmet need, as currently, there are no FDA approved therapies for this disabling stage of MS. There are several ongoing trials of BTKi in for this indication, and a recently completed phase 3 clinical trial of tolebrutinib in naSPMS (HERCULES) demonstrated a 31% delay in time to clinical progression however 4.1% experienced liver function test abnormalities with one death to hepatic toxicity.^57^ The effect on microglial activation in vivo was not measured. Moreover, the GEMINI 1 and 2 studies of tolebrutinib in RRMS failed to show a reduction in relapses compared to teriflunomide and tolebrutinib had a higher accumulation of Gd+ lesions.^58^ Thus, there is need for additional, novel approaches to SPMS.

In summary, nasal foralumab in treatment-refractory non-active SPMS patients showed stabilization in EDSS, improvement in fatigue scores and improvement in TSPO-PET imaging. This was accompanied by an increase in TGFΒ1 in monocytes amongst other immunological changes starting as early as three months after treatment. These results form the basis for an ongoing phase 2a multi-center randomized double-blind study comparing nasal foralumab to placebo (NCT06292923).

## Data Availability

All data produced in the present study are available upon reasonable request to the authors

## Acknowledgements

We thank the clinical research staff at the Translational Neuroimmunology Research Center and clinicians at the Brigham MS Center for their support. We thank Mariann Polgar-Turcsanyi for database management. The project’s resources were provided in part by the Center for Clinical Investigation (CCI) were supported by grant UL1TR002541(NIH/NCATS).

## Supplementary figures

**Supplementary Figure 1:** Time on foralumab treatment: Expanded Access patients treatment cycles depicted in diamonds. Straight lines indicate periods of treatment hiatus.

**Supplementary Figure 2A.** CSF Oligoclonal band testing at baseline and 6 months after nasal foralumab treatment: demonstrate change in CSF oligoclonal bands 6 months after nasal foralumab treatment; p=0.362, Paired t-test.

**Supplementary Figure 2B.** Olink 48-plex cytokine analysis in the CSF of foralumab-treated patients at baseline (T1 and 6 months (T2) shows a trend in reduction of CXCL12 (0.052).

**Supplementary Figure 3.** Standardized uptake value (SUV) maps showing a diffusely reduced [F-18]PBR06-PET brain uptake after 3 months (bottom row) of treatment with nasal foralumab in subject EA1, as compared to baseline (top row)

**Supplementary Figure 4. 4A**. Standardized uptake value (SUV) maps showing a diffusely reduced [F-18]PBR06-PET brain uptake after 3 months (middle row) and 6 months (bottom row) of treatment with nasal foralumab in other high affinity binders (EA3, EA4, EA6), as compared to baseline (top row). **4B**. Individualized mGALP z-score maps for high affinity binders (subjects EA6, EA1, EA4) superimposed on respective subject’s T2-FLAIR MRIs demonstrate decreased PET abnormalities following treatment at 3 months (middle row) and 6 months (bottom row) as compared to baseline (top row). Hotter colors represent a higher z-score value and cooler colors represent a lower z-score value as determined by the GALP 3.0 pipeline.

**Supplementary Figure 5.** Correlation between individualized, average voxel-by-voxel mGALP (GALP 3.0) z-scores in the hippocampus with overall modified fatigue impact scale (MFIS) score.

**Supplementary Figure 6.** Decreased hippocampal and substantia nigra PET signal in subjects with total MFIS>30 or cognitive MFIS>10 (n=5) after treatment with nasal foralumab. MFIS=Modified Fatigue Impact Scale.

**Supplementary Figure 7.** Individualized, voxel-by-voxel z-score maps focusing on lesional and perilesional white matter, superimposed on individual subjects’ T2 FLAIR MRI scans reveal evidence for reduced glial activity following **A**. 6 months and **B**. 3 months treatment with nasal foralumab in subjects EA1 and EA3 respectively.

## Contribution of each author

TC, TS and HLW contributed to the conception and design of the study, All authors contributed to the acquisition and analysis of data

TC, TS, BL, HLW contributed to drafting a significant portion of the manuscript or figures BL, BCH, HL, DK conducted statistical analysis

TC served as the study PI and IND holder for the foralumab protocol

TS served as IND holder and PI for the PET radiopharmaceutical and scans

## References

1. Kappos L, Wolinsky JS, Giovannoni G, et al. Contribution of Relapse-Independent Progression vs Relapse-Associated Worsening to Overall Confirmed Disability Accumulation in Typical Relapsing Multiple Sclerosis in a Pooled Analysis of 2 Randomized Clinical Trials. JAMA Neurol. 2020 Sep 1;77(9):1132–40.

2. Lublin FD, Haring DA, Ganjgahi H, et al. How patients with multiple sclerosis acquire disability. Brain. 2022 Sep 14;145(9):3147–61.

3. Chitnis T, Weiner HL. CNS inflammation and neurodegeneration. J Clin Invest. 2017 Oct 2;127(10):3577–87.

4. Kuhlmann T, Moccia M, Coetzee T, et al. Multiple sclerosis progression: time for a new mechanism-driven framework. Lancet Neurol. 2023 Jan;22(1):78–88.

5. Kutzelnigg A, Lucchinetti CF, Stadelmann C, et al. Cortical demyelination and diffuse white matter injury in multiple sclerosis. Brain. 2005 Nov;128(Pt 11):2705–12.

6. Weiner HL. Immune mechanisms and shared immune targets in neurodegenerative diseases. Nat Rev Neurol. 2025 Feb;21(2):67–85.

7. Cerovic V, Pabst O, Mowat AM. The renaissance of oral tolerance: merging tradition and new insights. Nat Rev Immunol. 2025 Jan;25(1):42–56.

8. Weiner HL, da Cunha AP, Quintana F, Wu H. Oral tolerance. Immunol Rev. 2011 May;241(1):241–59.

9. Rezende RM, Weiner HL. Cellular Components and Mechanisms of Oral Tolerance Induction. Crit Rev Immunol. 2018;38(3):207–31.

10. Wu HY, Quintana FJ, Weiner HL. Nasal anti-CD3 antibody ameliorates lupus by inducing an IL-10-secreting CD4+ CD25-LAP+ regulatory T cell and is associated with down-regulation of IL-17+ CD4+ ICOS+ CXCR5+ follicular helper T cells. J Immunol. 2008 Nov 1;181(9):6038–50.

11. Ochi H, Abraham M, Ishikawa H, et al. Oral CD3-specific antibody suppresses autoimmune encephalomyelitis by inducing CD4+ CD25-LAP+ T cells. Nat Med. 2006 Jun;12(6):627–35.

12. Kuhn C, Weiner HL. Therapeutic anti-CD3 monoclonal antibodies: from bench to bedside. Immunotherapy. 2016 Jul;8(8):889–906.

13. Ramos EL, Dayan CM, Chatenoud L, et al. Teplizumab and beta-Cell Function in Newly Diagnosed Type 1 Diabetes. N Engl J Med. 2023 Dec 7;389(23):2151–61.

14. Mayo L, Cunha AP, Madi A, et al. IL-10-dependent Tr1 cells attenuate astrocyte activation and ameliorate chronic central nervous system inflammation. Brain. 2016 Jul;139(Pt 7):1939–57.

15. Chitnis T, Kaskow BJ, Case J, et al. Nasal administration of anti-CD3 monoclonal antibody modulates effector CD8+ T cell function and induces a regulatory response in T cells in human subjects. Front Immunol. 2022;13:956907.

16. Van Camp N, Lavisse S, Roost P, Gubinelli F, Hillmer A, Boutin H. TSPO imaging in animal models of brain diseases. Eur J Nucl Med Mol Imaging. 2021 Dec;49(1):77–109.

17. Kreisl WC, Kim MJ, Coughlin JM, Henter ID, Owen DR, Innis RB. PET imaging of neuroinflammation in neurological disorders. Lancet Neurol. 2020 Nov;19(11):940–50.

18. Kaunzner UW, Kang Y, Zhang S, et al. Quantitative susceptibility mapping identifies inflammation in a subset of chronic multiple sclerosis lesions. Brain. 2019 Jan 1;142(1):133–45.

19. Nutma E, Fancy N, Weinert M, et al. Translocator protein is a marker of activated microglia in rodent models but not human neurodegenerative diseases. Nat Commun. 2023 Aug 28;14(1):5247.

20. Krasemann S, Madore C, Cialic R, et al. The TREM2-APOE Pathway Drives the Transcriptional Phenotype of Dysfunctional Microglia in Neurodegenerative Diseases. Immunity. 2017 Sep 19;47(3):566–81 e9.

21. Singhal T, Weiner HL, Bakshi R. TSPO-PET Imaging to Assess Cerebral Microglial Activation in Multiple Sclerosis. Semin Neurol. 2017 Oct;37(5):546–57.

22. Bodini B, Tonietto M, Airas L, Stankoff B. Positron emission tomography in multiple sclerosis - straight to the target. Nat Rev Neurol. 2021 Nov;17(11):663–75.

23. Singhal T, Cicero S, Pan H, et al. Regional microglial activation in the substantia nigra is linked with fatigue in MS. Neurol Neuroimmunol Neuroinflamm. 2020 Sep 3;7(5).

24. Sucksdorff M, Matilainen M, Tuisku J, et al. Brain TSPO-PET predicts later disease progression independent of relapses in multiple sclerosis. Brain. 2020 Dec 5;143(11):3318–30.

25. Laaksonen S, Saraste M, Sucksdorff M, et al. Early prognosticators of later TSPO-PET-measurable microglial activation in multiple sclerosis. Mult Scler Relat Disord. 2023 Jul;75:104755.

26. Singhal T, O’Connor K, Dubey S, et al. Gray matter microglial activation in relapsing vs progressive MS: A [F-18]PBR06-PET study. Neurol Neuroimmunol Neuroinflamm. 2019 Sep;6(5):e587.

27. Rissanen E, Tuisku J, Rokka J, et al. In Vivo Detection of Diffuse Inflammation in Secondary Progressive Multiple Sclerosis Using PET Imaging and the Radioligand (1)(1)C-PK11195. J Nucl Med. 2014 Jun;55(6):939–44.

28. Singhal T, Rissanen E, Ficke J, Cicero S, Carter K, Weiner HL. Widespread Glial Activation in Primary Progressive Multiple Sclerosis Revealed by 18F-PBR06 PET: A Clinically Feasible, Individualized Approach. Clin Nucl Med. 2021 Feb 1;46(2):136–7.

29. Singhal T, Cicero S, Rissanen E, et al. Glial Activity Load on PET Reveals Persistent “Smoldering” Inflammation in MS Despite Disease-Modifying Treatment: 18 F-PBR06 Study. Clin Nucl Med. 2024 Jun 1;49(6):491–9.

30. Albrecht DS, Mainero C, Ichijo E, et al. Imaging of neuroinflammation in migraine with aura: A [(11)C]PBR28 PET/MRI study. Neurology. 2019 Apr 23;92(17):e2038–e50.

31. Minoshima S, Frey KA, Koeppe RA, Foster NL, Kuhl DE. A diagnostic approach in Alzheimer’s disease using three-dimensional stereotactic surface projections of fluorine-18-FDG PET. J Nucl Med. 1995 Jul;36(7):1238–48.

32. Meier DS, Guttmann CRG, Tummala S, et al. Dual-Sensitivity Multiple Sclerosis Lesion and CSF Segmentation for Multichannel 3T Brain MRI. J Neuroimaging. 2018 Jan;28(1):36–47.

33. Zurawski J, Glanz BI, Healy BC, et al. The impact of cervical spinal cord atrophy on quality of life in multiple sclerosis. J Neurol Sci. 2019 Aug 15;403:38–43.

34. Barro C, Healy BC, Liu Y, et al. Serum GFAP and NfL Levels Differentiate Subsequent Progression and Disease Activity in Patients With Progressive Multiple Sclerosis. Neurol Neuroimmunol Neuroinflamm. 2023 Jan;10(1).

35. Hao Y, Stuart T, Kowalski MH, et al. Dictionary learning for integrative, multimodal and scalable single-cell analysis. Nat Biotechnol. 2024 Feb;42(2):293–304.

36. Satija R, Farrell JA, Gennert D, Schier AF, Regev A. Spatial reconstruction of single-cell gene expression data. Nat Biotechnol. 2015 May;33(5):495–502.

37. Stuart T, Butler A, Hoffman P, et al. Comprehensive Integration of Single-Cell Data. Cell. 2019 Jun 13;177(7):1888–902 e21.

38. Korsunsky I, Millard N, Fan J, et al. Fast, sensitive and accurate integration of single-cell data with Harmony. Nat Methods. 2019 Dec;16(12):1289–96.

39. Heaton H, Talman AM, Knights A, et al. Souporcell: robust clustering of single-cell RNA-seq data by genotype without reference genotypes. Nat Methods. 2020 Jun;17(6):615–20.

40. Hao Y, Hao S, Andersen-Nissen E, et al. Integrated analysis of multimodal single-cell data. Cell. 2021 Jun 24;184(13):3573–87 e29.

41. Love MI, Huber W, Anders S. Moderated estimation of fold change and dispersion for RNA-seq data with DESeq2. Genome Biol. 2014;15(12):550.

42. Liberzon A, Birger C, Thorvaldsdottir H, Ghandi M, Mesirov JP, Tamayo P. The Molecular Signatures Database (MSigDB) hallmark gene set collection. Cell Syst. 2015 Dec 23;1(6):417–25.

43. Rooney S, McFadyen DA, Wood DL, Moffat DF, Paul PL. Minimally important difference of the fatigue severity scale and modified fatigue impact scale in people with multiple sclerosis. Mult Scler Relat Disord. 2019 Oct;35:158–63.

44. Margoni M, Valsasina P, Bacchetti A, et al. Resting state functional connectivity modifications in monoaminergic circuits underpin fatigue development in patients with multiple sclerosis. Mol Psychiatry. 2024 Sep;29(9):2647–56.

45. Cicero S, Hansel C, Dubey S, et al. Cortical Microglial Activation Mediates the Relationship Between Fatigue and Progression Independent of Relapse Activity (PIRA) in Multiple Sclerosis: A 4.5-year Longitudinal Study. Americas Committee for Treatment and Research in Multiple Sclerosis (ACTRIMS); Februray 27, 2025 to March 1, 2025; West Palm Beach, Florida2025.

46. Sucksdorff M, Tuisku J, Matilainen M, et al. Natalizumab treatment reduces microglial activation in the white matter of the MS brain. Neurol Neuroimmunol Neuroinflamm. 2019 Jul;6(4):e574.

47. Sucksdorff M, Rissanen E, Tuisku J, et al. Evaluation of the Effect of Fingolimod Treatment on Microglial Activation Using Serial PET Imaging in Multiple Sclerosis. J Nucl Med. 2017 Oct;58(10):1646–51.

48. Ratchford JN, Endres CJ, Hammoud DA, et al. Decreased microglial activation in MS patients treated with glatiramer acetate. J Neurol. 2012 Jun;259(6):1199–205.

49. 49. Singhal T, Cicero S, Gale S, et al. Dampening of microglial activation with nasal foralumab administration in moderate Alzheimer’s Disease dementia. Clinical Nuclear Medicine (In press). 2025.

50. Lopes JR, Zhang X, Mayrink J, et al. Nasal administration of anti-CD3 monoclonal antibody ameliorates disease in a mouse model of Alzheimer’s disease. Proc Natl Acad Sci U S A. 2023 Sep 12;120(37):e2309221120.

51. Izzy S, Yahya T, Albastaki O, et al. Nasal anti-CD3 monoclonal antibody ameliorates traumatic brain injury, enhances microglial phagocytosis and reduces neuroinflammation via IL-10-dependent T(reg)-microglia crosstalk. Nat Neurosci. 2025 Feb 27.

52. Kuhn C, Rezende RM, da Cunha AP, et al. Mucosal administration of CD3-specific monoclonal antibody inhibits diabetes in NOD mice and in a preclinical mouse model transgenic for the CD3 epsilon chain. J Autoimmun. 2017 Jan;76:115–22.

53. Boden EK, Canavan JB, Moran CJ, et al. Immunologic Alterations Associated With Oral Delivery of Anti-CD3 (OKT3) Monoclonal Antibodies in Patients With Moderate-to-Severe Ulcerative Colitis. Crohns Colitis 360. 2019 Jul;1(2):otz009.

54. Moreira TG, Matos KTF, De Paula GS, et al. Nasal Administration of Anti-CD3 Monoclonal Antibody (Foralumab) Reduces Lung Inflammation and Blood Inflammatory Biomarkers in Mild to Moderate COVID-19 Patients: A Pilot Study. Front Immunol. 2021;12:709861.

55. Sims EK, Bundy BN, Stier K, et al. Teplizumab improves and stabilizes beta cell function in antibody-positive high-risk individuals. Sci Transl Med. 2021 Mar 3;13(583).

56. Carr MW, Roth SJ, Luther E, Rose SS, Springer TA. Monocyte chemoattractant protein 1 acts as a T-lymphocyte chemoattractant. Proc Natl Acad Sci U S A. 1994 Apr 26;91(9):3652–6.

57. Fox RJ, Bar-Or A, Traboulsee A, et al. Tolebrutinib in Nonrelapsing Secondary Progressive Multiple Sclerosis. N Engl J Med. 2025 Apr 8.

58. Oh J, Arnold DL, Cree BAC, et al. Tolebrutinib versus Teriflunomide in Relapsing Multiple Sclerosis. N Engl J Med. 2025 Apr 8.

