## Supplemental tables and figures for "Nasal foralumab treatment of PIRA induces regulatory immunity, dampens microglial activation and stabilizes clinical progression in non-active secondary progressive MS"

### Supplementary materials

| | Baseline | Six month | Change | Improvement |
|---|---|---|---|---|
| **T25FW** | 12.6 (7.9) | 14.4 (10.6) | 1.8 (4.4) p=0.16 | 1/10 |
| **9-HPT** | 28.5 (5.9) | 30.9 (9.9) | 2.3 (6.1) p=0.44 | 0/10 |
| **SDMT** | 40.6 (10.7) | 47.7 (8.5) | 7.1 (4.3) p=0.009 | 9/10 |
| **LCLA*** | 23.3 (14.9) | 30.8 (15.0) | 6.6 (7.9) p=0.049 | 4/9 |

*There was one patient missing the LCLA at the six month time point.

Supplementary Table 1: MSFC4 scores at baseline and 6 months in foralumab-treated patients.

| | Baseline | 6 months | Change | Wilcoxon p-value |
|---|---|---|---|---|
| **Anxiety** | 43.91 (11.68) n=10 | 41.02 (6.44) n=8 | -1.12 (9.91) n=8 | 0.5896 |
| **Depression** | 41.65 (4.23) n=10 | 40.02 (6.2) n=9 | -1.22 (5.51) n=9 | 0.4164 |
| **Fatigue** | 48.04 (4.2) n=9 | 48.08 (10.12) n=10 | -0.9 (6.66) n=9 | 0.6523 |
| **Upper** | 43.84 (7.54) n=10 | 43.09 (6.79) n=10 | -0.75 (7.77) n=10 | 0.8885 |
| **Lower** | 40.36 (5.64) n=10 | 40.99 (7.62) n=10 | 0.63 (4.17) n=10 | 0.6953 |
| **Cognitive** | 53.66 (6.65) n=9 | 57.96 (7.49) n=10 | 3.61 (4.21) n=9 | 0.036 |
| **Emotional** | 43.66 (5.88) n=10 | 40.8 (7.33) n=9 | -2.52 (3.81) n=9 | 0.0759 |
| **PositiveAffect** | 56.82 (5.03) n=9 | 60.84 (5.9) n=10 | 3.74 (6.37) n=9 | 0.1083 |
| **Sleep** | 45.97 (7.92) n=9 | 41.88 (11.96) n=10 | -2.99 (14.58) n=9 | 0.9326 |
| **Social** | 47.6 (7.14) n=10 | 48.69 (8.54) n=8 | 0.65 (6.7) n=8 | 0.8551 |
| **Satisfaction** | 43.73 (2.44) n=10 | 44.29 (5.48) n=10 | 0.56 (3.73) n=10 | 0.6953 |
| **Stigma** | 51.17 (4.39) n=10 | 47.45 (14.32) n=10 | -3.72 (13.66) n=10 | 1 |

Supplementary Table 2: NeuroQOL short subscales demonstrate no significant change at 6 months compared to baseline.

Supplementary Table 3: mGALP scores for baseline, 3 months and 6 months on treatment

| | **Baseline** | **3-month** | **Change** | **Wilcoxon signed rank p-value** |
|---|---|---|---|---|
| Group | 0.42 (0.36) | 0.31 (0.23) | -0.11 (0.14) | 0.016 |
| Cortex | 0.37 (0.47) | 0.25 (0.28) | -0.12 (0.20) | 0.11 |
| Thalamus | 0.99 (0.90) | 0.84 (0.61) | -0.15 (0.44) | 0.46 |
| White matter | 0.47 (0.09) | 0.40 (0.09) | -0.07 (0.08) | 0.055 |
| Cerebellum | 0.49 (0.60) | 0.33 (0.57) | -0.16 (0.20) | 0.11 |
| | **Baseline** | **6-month*** | **Change*** | **Wilcoxon signed rank p-value*** |
| Group | 0.42 (0.36) | 0.27 (0.24) | -0.11 (0.15) | 0.30 |
| Cortex | 0.37 (0.47) | 0.19 (0.29) | -0.12 (0.21) | 0.30 |
| Thalamus | 0.99 (0.90) | 0.84 (0.67) | -0.14 (0.87) | 0.69 |
| White matter | 0.47 (0.09) | 0.40 (0.08) | -0.07 (0.08) | 0.078 |
| Cerebellum | 0.49 (0.60) | 0.25 (0.55) | -0.20 (0.26) | 0.11 |

*Legend: *One subject was removed from the 6-month and 6-month change analysis*

Supplementary Table 6

### APPENDIX 1: DETAILED SCHEDULE OF EVENTS FOR NEW PATIENTS

| Activity | Screening[1] | Dosing | Rest | Dosing | Rest | Dosing | Rest | Dosing | Rest | Dosing | Rest | Dosing | Rest | Dosing | Rest | Dosing/EOT | Rest |
|---|---|---|---|---|---|---|---|---|---|---|---|---|---|---|---|---|---|
| **Cycle Number** | 0 | 1 | | 2 | | 3 | | 4 | | 5 | | 6 | | 7 | | 8[10] | |
| **Cycle Week** | 0 | 1-2 | 3 | 1-2 | 3 | 1-2 | 3 | 1-2 | 3 | 1-2 | 3 | 1-2 | 3 | 1-2 | 3 | 1-2 | 3 |
| **Cycle Day** | 0 | 1-14 | 15-21 | 1-14 | 15-21 | 1-14 | 15-21 | 1-14 | 15-21 | 1-14 | 15-21 | 1-14 | 15-21 | 1-14 | 15-21 | 1-14 | 15-21 |
| **Day** | -14-0 | 1-14 | 15-21 | 22-35 | 36-42 | 43-56 | 57-63 | 64-77 | 78-84 | 85-98 | 99-105 | 106-119 | 120-126 | 127-139 | 140-146 | 147-160 | 161-167 |
| **Informed Consent** | X | | | | | | | | | | | | | | | | |
| **Medical History & Demography** | X | X | | X | | X | | X | | X | | X | | X | | X | |
| **Concomitant Meds** | X | X | | X | | X | | X | | X | | X | | X | | X | |
| **Review of Adverse Events** | | X | | X | | X | | X | | X | | X | | X | | X | |
| **Physical Exam[2]** | X | X | | X | | X | | X | | X | | X | | X | | X | |
| **Neurologic Exam[3] (Detailed)** | X | X | | X | | X | | X | | X | | X | | X | | X | |
| **Nasal Exam by ENT[4]** | X | | | | | | | X | | | | | | | | X | |
| **Hematology & Chemistry** | X | X | | X | | X | | X | | X | | X | | X | | X | |
| **Immunology & ADAs** | X | X | | X | | X | | X | | X | | X | | X | | X | |
| **NSQ[5]** | X | X | | X | | X | | X | | X | | X | | X | | X | |
| **Vitals Signs[5]** | X | X | | X | | X | | X | | X | | X | | X | | X | |
| **In-Clinic Dosing, Each Day 1[6]** | | X | | X | | X | | X | | X | | X | | X | | X | |
| **Monitoring Post-Dose[7]** | | X[7] | | X | | X | | X | | X | | X | | X | | X | |
| **Dispense At-Home IP** | | X | | X | | X | | X | | X | | X | | X | | X | |
| **At-Home Dosing[8]** | | X | | X | | X | | X | | X | | X | | X | | X | |
| **MSFC-4, MFIS, CVLT-2** | X | X | | X | | X | | X | | X | | X | | X | | X | |
| **NeuroQoL** | X | X | | | | X | | | | X | | | | X | | X | |
| **ECG** | X | | | | | | | X | | | | | | | | X | |
| **Lumbar Puncture (LP)** | X | | | | | | | | | | | | | | | X | |
| **PET scan** | X | | | | | | | X[9] | | | | | | | | X[9] | |
| **3T MRI scan, brain/spine (incl QSM)** | X | | | | | | | X[9] | | | | | | | | X[9] | |
| **7T MRI scan** | X | | | | | | | | | | | | | | | X[9] | |

1. Activities may take place over multiple visits; consent will always be obtained first. Physical exams conducted during screening within 7 days of Cycle 1 do not need to be reperformed.
2. Physical exam and labs should be done on Day 1 of each cycle but may be conducted during the rest week or on Day 3 of each cycle, always prior to dosing.
3. Neurologic Exam includes EDSS score.
4. Nasal exam must occur no later than the second dosing day in the first week of each cycle, always prior to dosing or during the rest week.
5. NSQ administered and vitals obtained twice during dosing visits, pre- and post-dose (1 hour following dosing).
6. In-clinic dosing is required Day 1 of each cycle to ensure patient is capable of self-administration and to review previous cycle's diary and NSQs. At-home dosing will occur following Day 1 visit.
7. Monitoring includes vital signs and only occurs after dosing occurring in-clinic. Cycle 1, day 1 monitoring is 2 hours and prospective day 1 dosing is 1 hour.
8. At-home dosing will occur on either on Wednesday and Friday or Thursday and Saturday during week 1, and Monday, Wednesday, Friday during week 2.
9. Cycle 4 and 8 imaging may occur during the respective treatment cycle and should occur prior to the proceeding treatment cycle whenever possible.
10. Unscheduled or early termination visits will follow Cycle 8 procedures. Cycle 8 activities may take place over multiple visits. For patients not continuing treatment beyond 6 months, Cycle 8 visit will be their final visit.

EOT = End of treatment
Hematology = CBC with differential
Chemistry = includes urinalysis; Na, K, glucose, CO2, Total bilirubin, Direct bilirubin, Creatinine, BUN, EBV serology
Immunology = 1 red top, 1 purple top, 5 green top tubes
NSQ = Nasal Symptom Questionnaire

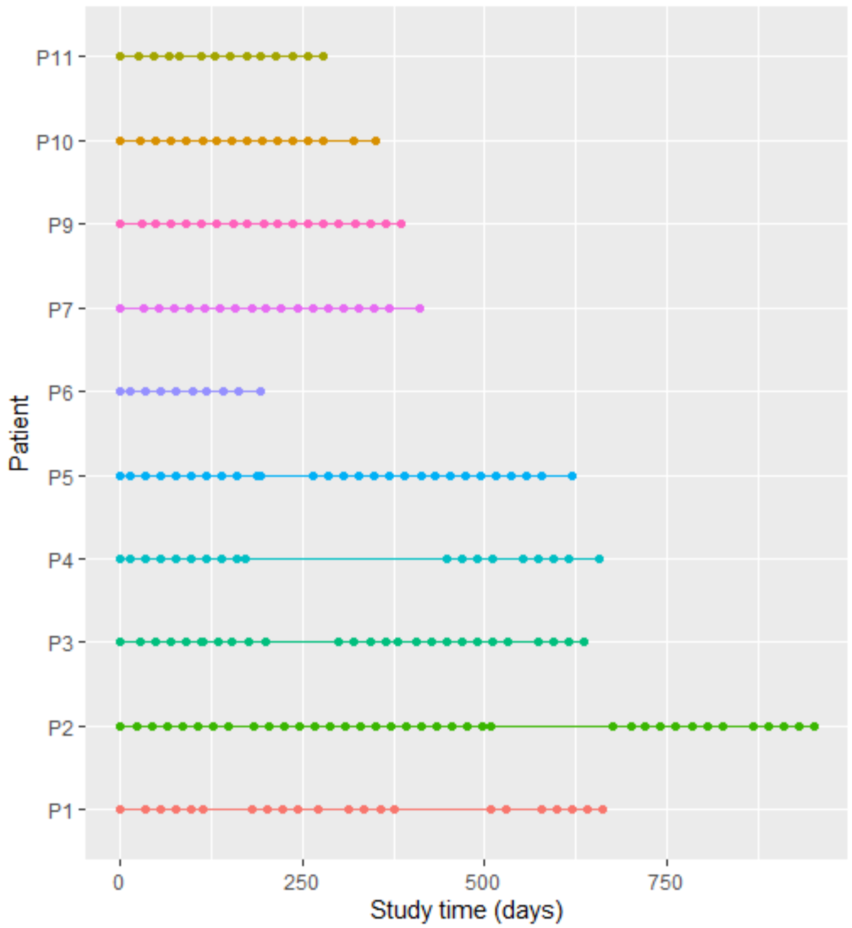


Supplementary Figure 1

#### CSF Oligoclonal bands at baseline and 6 mos after foralumab treatment

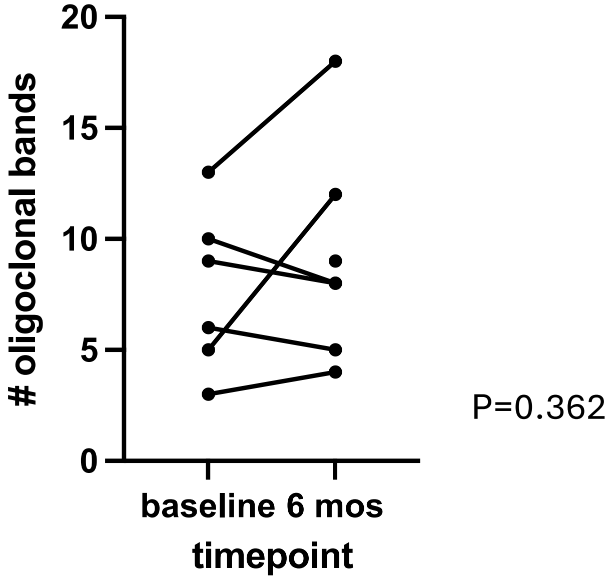


Supplementary Figure 2a

Supplementary Figure 2b

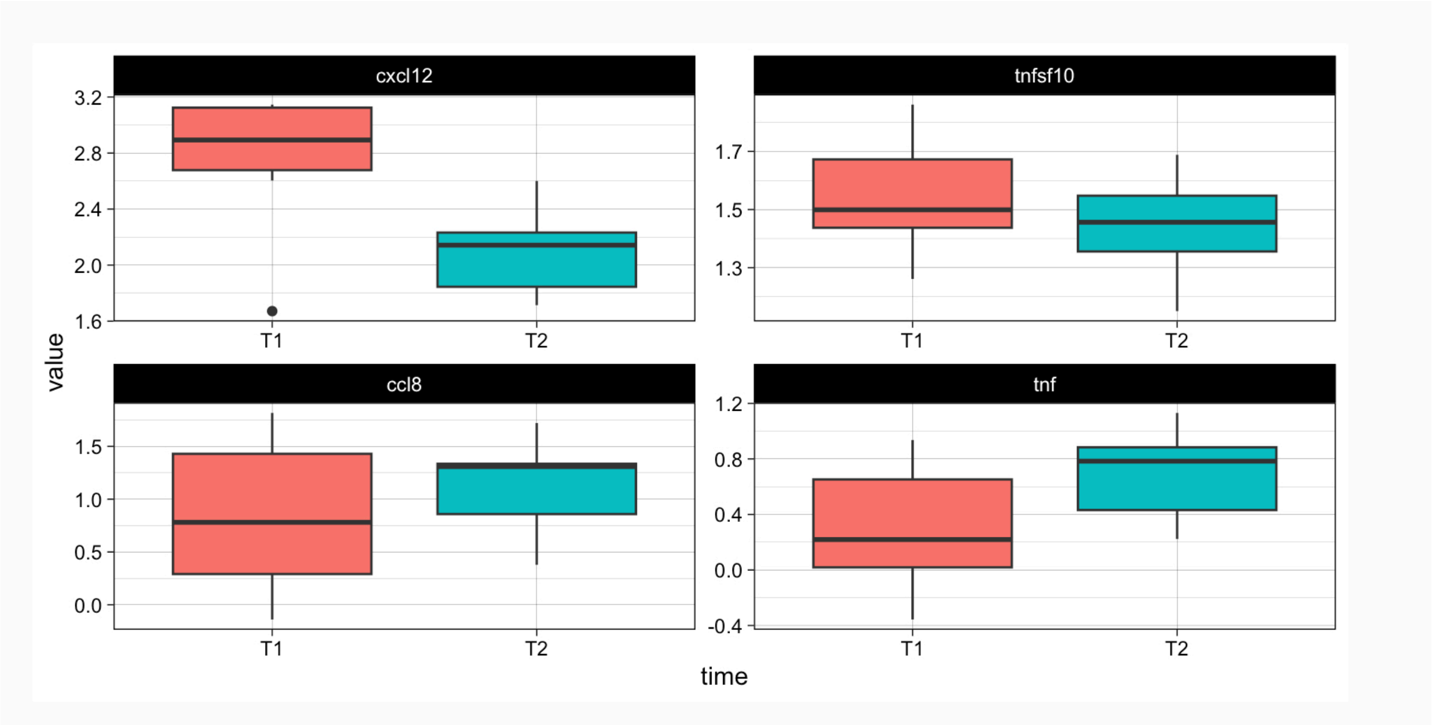

SUPPL FIG 3

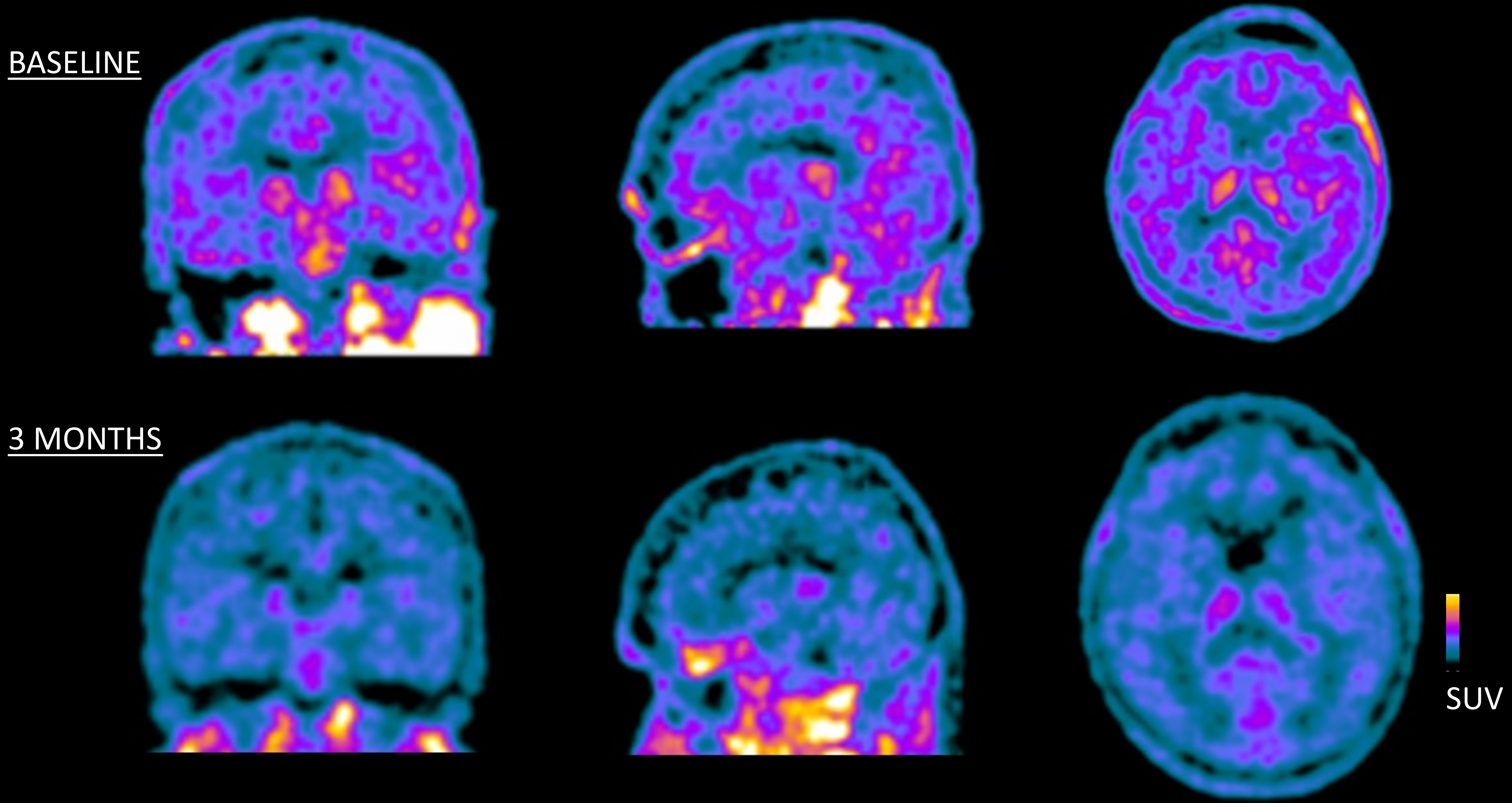

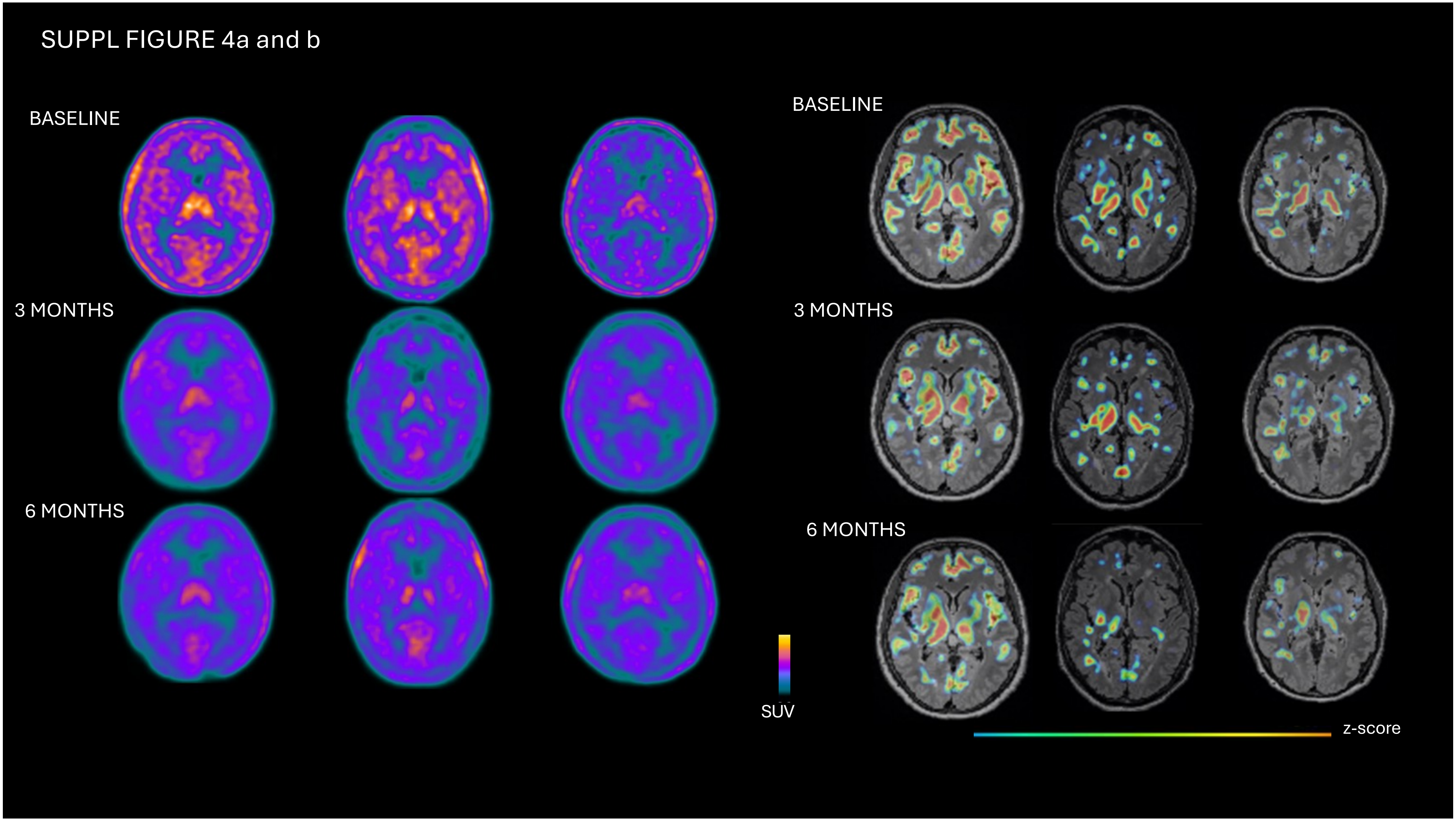
SUPPL FIGURE 4a and b
BASELINE
3 MONTHS
6 MONTHS
SUV
BASELINE
3 MONTHS
6 MONTHS
z-score

SUPPL FIGURE 5: Correlation of fatigue scores measured with MFIS with TSPO-PET Hippocampal signal

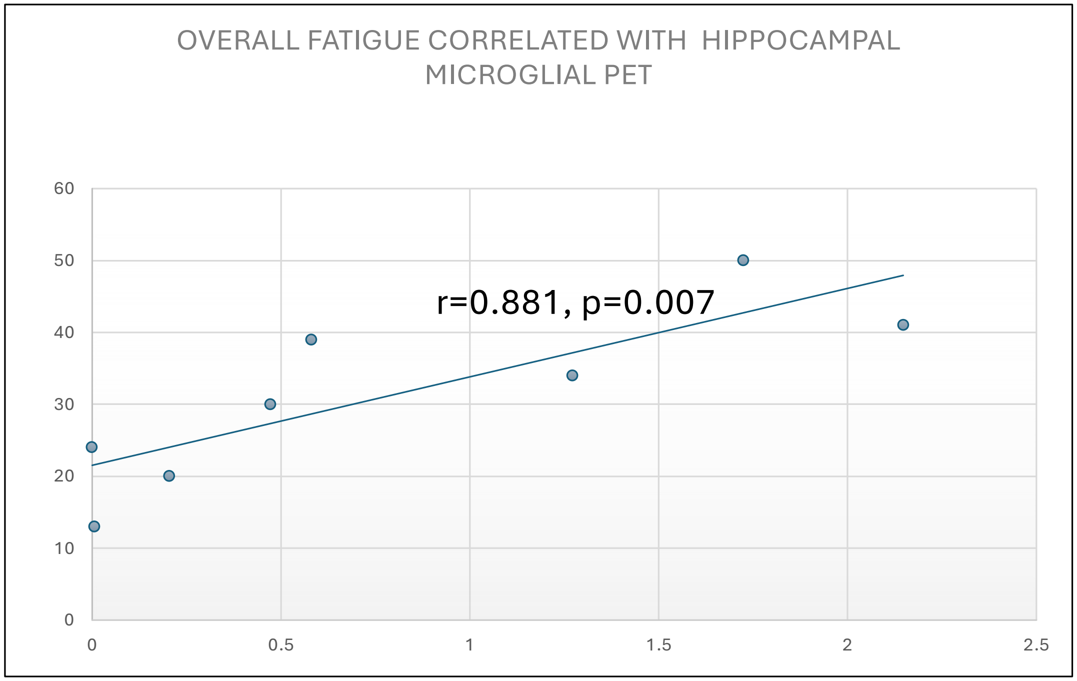

Supplementary Figure 6

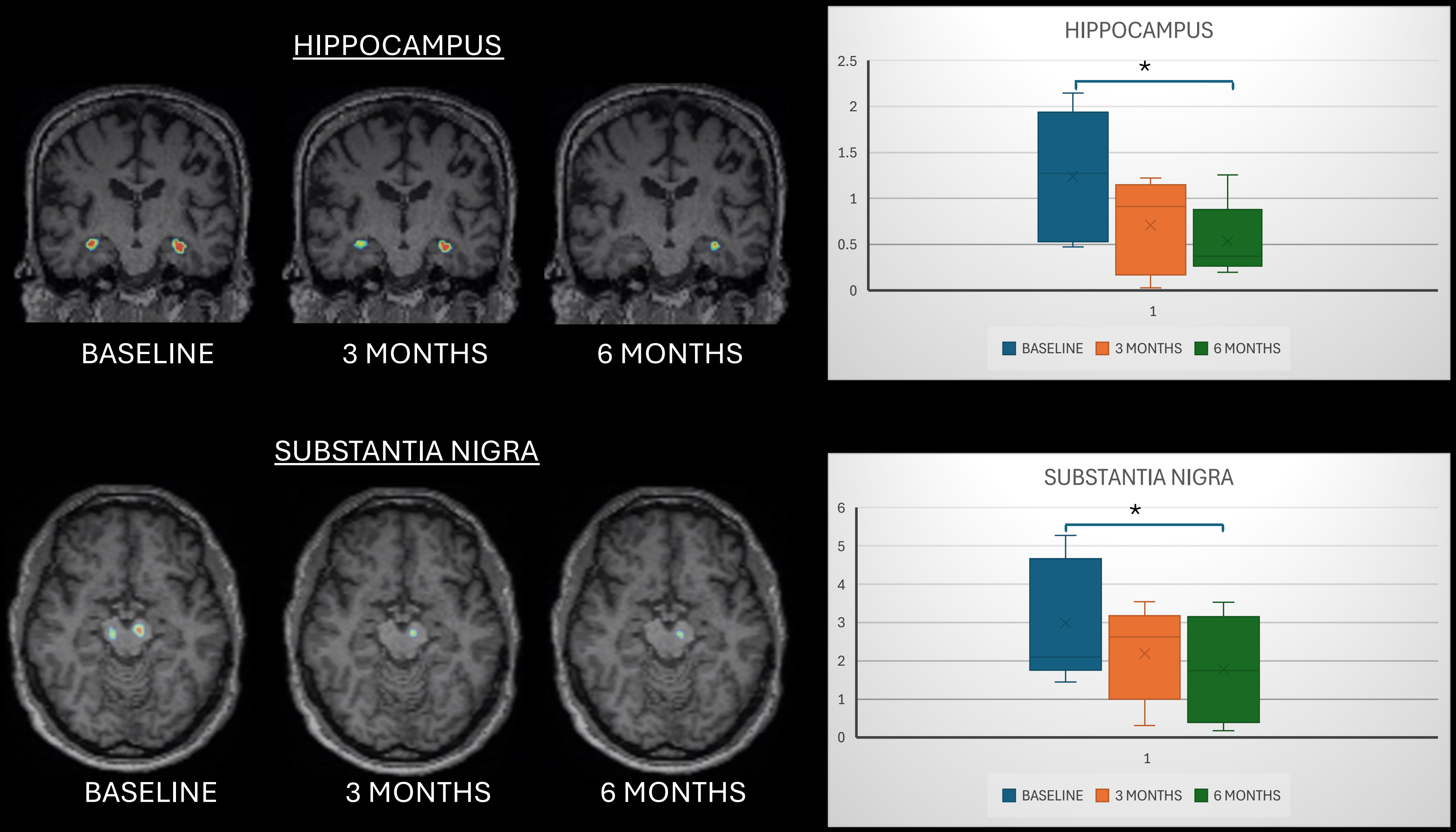

SUPPL Figure 7a and 7b

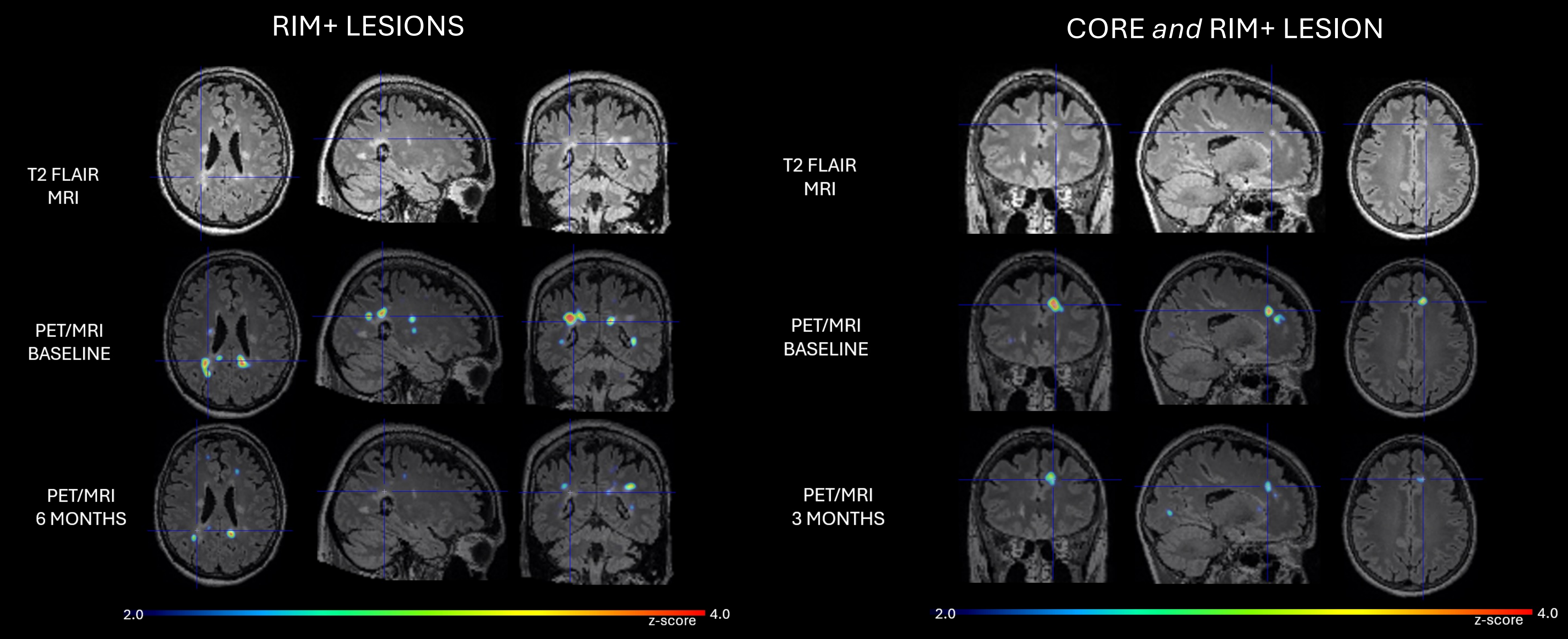
