## Supplemental Table 4 for "Nasal foralumab treatment of PIRA induces regulatory immunity, dampens microglial activation and stabilizes clinical progression in non-active secondary progressive MS"

Supplementary Table 4:	Incidence of Treatment Emergent Adverse Events (TEAEs)

| **Adverse Event** | **Subject ID(s)** | **Relatedness** | **Mild*** | **Moderate*** | **Severe*** | **Total**** |
| --- | --- | --- | --- | --- | --- | --- |
| Abnormal lab value – alkaline phosphate | EA7 | UR |  | 1 |  | 1 |
| Abnormal lab value – high ALT | EA11, EA12 | UR |  | 1 | 2 | 3 |
| Abnormal lab value – high bilirubin | EA9, EA11, EA12 | UR |  | 3 |  | 3 |
| Abnormal lab value – high BUN | EA4, EA10 | UR |  | 2 | 1 | 3 |
| Abnormal lab value – high leukocytes | EA9 | UR |  | 1 |  | 1 |
| Abnormal lab value – high lymphocytes | EA13 | UR |  | 1 |  | 1 |
| Abnormal lab value – high monocytes | EA12 | UR |  | 1 |  | 1 |
| Abnormal lab value – high RBCs | EA5, EA7, EA11 | UR |  | 3 | 1 | 4 |
| Abnormal lab value – high WBCs | EA7, EA9 | UR |  | 2 | 2 | 4 |
| Abnormal lab value – hyperglycemia | EA4, EA10, EA11, EA15 | UR |  | 5 |  | 5 |
| Abnormal lab value – hypernatremia | EA11 | UR |  | 2 |  | 2 |
| Abnormal lab value – hypoglycemia | EA2, EA3, EA5, EA9, EA10 | UR |  | 7 | 3 | 10 |
| Abnormal lab value – hyponatremia | EA5, EA9 | UR |  |  | 3 | 3 |
| Abnormal lab value – leukocyte esterase | EA5, EA7, EA13 | UR |  | 5 | 1 | 6 |
| Abnormal lab value – low hemoglobin | EA2, EA5 | UR |  | 2 |  | 2 |
| Abnormal lab value – low lymphocytes | EA4 | UR |  | 1 |  | 1 |
| Abnormal lab value – urine nitrites | EA9 | UR |  | 1 |  | 1 |
| Abnormal lab value – urine proteins | EA7 | UR |  | 1 |  | 1 |
| Abnormal nasal exam – scant blood | EA2 | PR |  | 1 |  | 1 |
| Abnormal nasal exam – scant blood, anterior septum | EA2 | PR |  | 1 |  | 1 |
| Abnormal nasal exam – erythema crusting and blood in the anterior nasal cavity (septum, inferior turbinate, nasal sill) | EA6 | UR | 1 |  |  | 1 |
| Abnormal nasal exam – inferior turbinate moderate hypertrophy, interior head left middle turbinate with trace blood - associated with surgery | EA5 | PR | 1 |  |  | 1 |
| Abnormal nasal exam – inferior turbinate severe hypertrophy (right), inferior turbinate moderate to severe hypertrophy (left), scant bleeding on right septum | EA5 | PR |  | 1 |  | 1 |
| Abnormal nasal exam – mild bilateral nasal vestibulitis and blood/scabbing (left) and scant mucus drainage | EA3 | UR | 1 |  |  | 1 |
| Abnormal nasal exam – mild bilateral nasal vestibulitis, blood/scabbing, as well as moderate inferior turbinate hypertrophy bilaterally - associated with allergies | EA3 | PR | 1 |  |  | 1 |
| Abnormal nasal exam – mild hypertrophy (right), mild to moderate hypertrophy (left), and trace blood on septum - associated with allergies | EA6 | PR | 1 |  |  | 1 |
| Abnormal nasal exam – mild septal deviation, moderate to severe inferior turbinate hypertrophy, scant mucus on the left | EA5 | UR | 1 |  |  | 1 |
| Abnormal nasal exam – moderate to severe turbinate hypertrophy, moderate septal deviation - associated with infection | EA6 | UR |  | 1 |  | 1 |
| Abnormal nasal exam – scant moist crusting with 2mm area of fresh blood in anterior right septum - associated with allergies | EA7 | UR | 1 |  |  | 1 |
| Abrasions, leg | EA11 | UR |  | 1 |  | 1 |
| Aches, body | EA9 | UR | 2 |  |  | 2 |
| Aches, body – associated with infection or vaccine | EA2, EA3 | UR |  | 2 | 1 | 3 |
| Aches, hand | EA11 | UR | 1 |  |  | 1 |
| Bloody nose | EA3 | PR | 1 |  |  | 1 |
| Bloody nose | EA3, EA9 | UR | 2 |  |  | 2 |
| Blurry vision | EA5, EA6 | UR | 4 |  |  | 4 |
| Brain Fog | EA5, EA6, EA9 | UR | 1 | 2 |  | 3 |
| Broken tooth | EA6 | UR | 1 |  |  | 1 |
| Bruising, isolated areas – associated with motor vehicle accident | EA4 | UR | 2 |  | 2 | 4 |
| Burning with urination | EA5 | UR | 1 |  |  | 1 |
| Cancer, prostate | EA1 | UR |  | 1 |  | 1 |
| Chills – associated with infection or vaccine | EA1, EA2, EA3 | UR |  | 2 | 2 | 4 |
| Cold sore | EA9 | UR | 1 |  |  | 1 |
| Congestion, nasal | EA5, EA6 | PR | 1 | 7 |  | 8 |
| Congestion, nasal | EA5, EA6, EA7, EA9, EA11 | UR | 4 | 1 |  | 5 |
| Congestion, nasal – associated with infection or vaccine | EA1 | UR |  | 1 |  | 1 |
| Congestion, respiratory | EA1 |  |  | 1 |  | 1 |
| Constipation | EA2, EA92 | UR | 2 |  |  | 2 |
| Contact dermatitis | EA10 | UR | 1 |  |  | 1 |
| Corneal scratches | EA5 | UR | 1 |  |  | 1 |
| Cough | EA3, EA5, EA7 | UR | 2 | 1 |  | 3 |
| Cough – associated with infection or vaccine | EA3 | UR |  | 1 |  | 1 |
| COVID-19 infection | EA1, EA5, EA9, EA7, EA11 | UR | 1 | 5 | 3 | 9 |
| Diarrhea | EA1 | PR |  | 1 |  | 1 |
| Diarrhea | EA3 | UR | 1 |  |  | 1 |
| Difficulty with sleep | EA6 | PR |  | 1 |  | 1 |
| Discomfort, isolated areas | EA3, EA6 | UR | 2 |  |  | 2 |
| Dizziness | EA5 | UR | 1 |  |  | 1 |
| Dryness, eye | EA5 | UR | 1 |  |  | 1 |
| Edema, lower extremities | EA6 | UR | 2 |  |  | 2 |
| Fall | EA6 | PR |  | 1 |  | 1 |
| Fall | EA3, EA4, EA6, EA11 | UR | 2 | 5 |  | 7 |
| Fall, repeated | EA11 | UR |  | 7 |  | 7 |
| Fatigue | EA9 | PR | 1 |  |  | 1 |
| Fatigue | EA1, EA5, EA9 | UR | 1 | 2 |  | 3 |
| Fatigue – associated with infection or vaccine | EA2, EA3, EA4, EA9 | UR | 1 | 3 | 1 | 5 |
| Fever | EA2, EA9 | UR | 3 |  |  | 3 |
| Fever – associated with infection or vaccine | EA1, EA2, EA7 | UR | 1 | 1 | 2 | 4 |
| Flatulence | EA9 | UR | 1 |  |  | 1 |
| Headache | EA2, EA10 | PR |  | 3 |  | 3 |
| Headache | EA4, EA5, EA6, EA9, EA11, EA15 | UR | 11 | 2 |  | 13 |
| Headache – associated with infection, vaccine or motor vehicle accident | EA1, EA3, EA4 | UR | 1 | 2 | 1 | 4 |
| Heart racing | EA5 | UR | 1 |  |  | 1 |
| Hypertension | EA2 | UR |  |  | 1 | 1 |
| Irritation, GI | EA12 | UR | 2 |  |  | 2 |
| Itching, nasal | EA1 | PR | 1 |  |  | 1 |
| Laceration – associated with motor vehicle accident | EA4 | UR |  |  | 1 | 1 |
| Lack of sensation, fingertips and legs | EA6 | UR | 1 |  |  | 1 |
| Lesion, shin | EA5 | UR | 1 |  |  | 1 |
| Lightheadedness | EA5 | PR | 1 |  |  | 1 |
| Lightheadedness | EA5 | UR | 1 |  |  | 1 |
| Memory loss – associated with motor vehicle accident | EA4 | UR |  |  | 1 | 1 |
| Mesenteric panniculitis – associated with motor vehicle accident | EA4 | UR |  |  | 1 | 1 |
| Mobility reduction – associated infection or vaccine | EA3, EA7 | UR |  | 4 |  | 4 |
| MS symptom worsening – associated infection or vaccine | EA2 | UR |  |  | 1 | 1 |
| Nausea | EA3 | UR | 1 |  |  | 1 |
| Nausea – associated with motor vehicle accident | EA4 | UR |  |  | 1 | 1 |
| Numbness, lips and tongue | EA9 | R | 1 |  |  | 1 |
| Numbness, isolated | EA5, EA9 | UR | 3 |  |  | 3 |
| Numbness, isolated – associated with surgery | EA5 | UR | 1 |  |  | 1 |
| Pain and pressure, sinus | EA9 | UR | 1 |  |  | 1 |
| Pain, isolated | EA3, EA5, EA6, EA9, EA11 | UR | 6 | 2 |  | 8 |
| Pain, isolated – associated with infection or vaccine, surgery or procedure, or motor vehicle accident | EA2, EA4, EA5 | UR | 1 | 2 | 3 | 6 |
| Pneumonia | EA2 | UR |  |  | 1 | 1 |
| Pneumothorax | EA2 | UR |  |  | 1 | 1 |
| Postnasal drip | EA5, EA7 | UR | 2 | 1 |  | 3 |
| Pressure with urination | EA9 | UR | 1 |  |  | 1 |
| Rash (poison ivy) | EA4 | UR | 1 |  |  | 1 |
| Runny Nose | EA1, EA5, EA6 | PR | 2 | 2 |  | 4 |
| Runny Nose | EA4, EA5, EA7 | UR | 3 |  |  | 3 |
| Runny nose – associated infection or vaccine | EA3 | UR | 1 | 1 |  | 2 |
| Sneezing | EA9 | UR | 1 |  |  | 1 |
| Sore throat | EA4, EA5, EA6 | UR | 3 |  |  | 3 |
| Sore throat – associated infection or vaccine | EA3, EA4 | UR | 1 | 1 |  | 2 |
| Soreness, isolated | EA4 | UR | 1 |  |  | 1 |
| Soreness, isolated – associated infection or vaccine | EA2 | UR | 1 |  |  | 1 |
| Spasms, back – associated infection or vaccine | EA1 | UR | 1 |  |  | 1 |
| Spasticity, transient | EA1 | PR |  | 1 |  | 1 |
| Spasticity, right leg | EA11 | UR |  | 1 |  | 1 |
| Stiffness, legs – associated infection or vaccine | EA7 | UR |  | 1 |  | 1 |
| Strain, lower back | EA9 | UR | 1 |  |  | 1 |
| Surgery or procedure – prostate hyperplasia | EA1 | UR | 1 |  |  | 1 |
| Surgery or procedure – tendon repair, quadriceps | EA11 | UR |  | 1 |  | 1 |
| Tachycardia | EA2 | UR |  |  | 1 | 1 |
| Tightness with breathing – associated infection or vaccine | EA2 | UR |  | 1 |  | 1 |
| Tightness, isolated | EA5 | UR | 2 |  |  | 2 |
| Tingling, lips and tongue | EA9 | R | 1 |  |  | 1 |
| Tingling, isolated | EA5 | UR | 1 |  |  | 1 |
| Toothache | EA10 | UR | 1 |  |  | 1 |
| Trigeminal neuralgia | EA5 | UR | 1 |  |  | 1 |
| Twitching, eye intermittent | EA6 | PR | 1 |  |  | 1 |
| Unsteadiness | EA6 | PR |  | 1 |  | 1 |
| Unsteadiness | EA6 | UR |  | 1 |  | 1 |
| Urinary tract infection | EA1, EA2, EA5, EA6, EA7, EA9 EA12 | UR | 11 | 4 | 3 | 18 |
| Vestibular issues, ears | EA5 | UR | 1 |  |  | 1 |
| Vomiting | EA3 | UR | 1 |  |  | 1 |
| Weakness, isolated | EA11 | UR |  | 1 |  | 1 |
| Weakness, isolated – associated infection or vaccine | EA4 | UR | 1 |  |  | 1 |
|  |  |  | 125 | 121 | 42 | 288 |

*Number of patients experiencing an adverse event (patient is to be counted only once for each adverse event)

**Total number of events

Severity: Mild, Moderate (Mod), Severe (Sev)

Relatedness: R = related; PR = possibly related; UR = unrelated

*Each participant is counted only once at the highest level of severity for the event. This table represents severity of all adverse events sorted in descending order of incidence as shown above; or adverse events related to the intervention as judged by the investigator; or treatment emergent event.*

*AEs determined from both nasal exam or lab results findings (noted as ‘nasal exam’ and ‘labs’).*
