## Supplemental Table 5 for "Nasal foralumab treatment of PIRA induces regulatory immunity, dampens microglial activation and stabilizes clinical progression in non-active secondary progressive MS"

Supplementary Table 5:	Incidence of Treatment Related Adverse Events (TRAE)

| **AE Term** | **Subject ID(s)** | **Relatedness** | **Mild*** | **Moderate*** | **Severe*** | **Total**** |
| --- | --- | --- | --- | --- | --- | --- |
| Abnormal nasal exam – scant blood | EA2 | PR |  | 1 |  | 1 |
| Abnormal nasal exam – scant blood, anterior septum | EA2 | PR |  | 1 |  | 1 |
| Abnormal nasal exam – inferior turbinate moderate hypertrophy, interior head left middle turbinate with trace blood - associated with surgery | EA5 | PR | 1 |  |  | 1 |
| Abnormal nasal exam – inferior turbinate severe hypertrophy (right), inferior turbinate moderate to severe hypertrophy (left), scant bleeding on right septum | EA5 | PR |  | 1 |  | 1 |
| Abnormal nasal exam – mild bilateral nasal vestibulitis, blood/scabbing, as well as moderate inferior turbinate hypertrophy bilaterally - associated with allergies | EA3 | PR | 1 |  |  | 1 |
| Abnormal nasal exam – mild hypertrophy (right), mild to moderate hypertrophy (left), and trace blood on septum - associated with allergies | EA6 | PR | 1 |  |  | 1 |
| Bloody nose | EA3 | PR | 1 |  |  | 1 |
| Congestion, nasal | EA5, EA6 | PR | 1 | 7 |  | 8 |
| Diarrhea | EA1 | PR |  | 1 |  | 1 |
| Difficulty with sleep | EA6 | PR |  | 1 |  | 1 |
| Fall | EA6 | PR |  | 1 |  | 1 |
| Fatigue | EA9 | PR | 1 |  |  | 1 |
| Headache | EA2, EA10 | PR |  | 3 |  | 3 |
| Lightheadedness | EA5 | PR | 1 |  |  | 1 |
| Numbness, lips and tongue | EA9 | R | 1 |  |  | 1 |
| Runny Nose | EA1, EA5, EA6 | PR | 2 | 2 |  | 4 |
| Spasticity, transient | EA1 | PR |  | 1 |  | 1 |
| Tingling, lips and tongue | EA9 | R | 1 |  |  | 1 |
| Twitching, eye intermittent | EA6 | PR | 1 |  |  | 1 |
| Unsteadiness | EA6 | PR |  | 1 |  | 1 |
|  |  |  | 15 | 18 | 0 | 33 |

*Number of patients experiencing an adverse event (patient is to be counted only once for each adverse event)

**Total number of events

Severity: Mild, Moderate (Mod), Severe (Sev)

Relatedness: R = related; PR = possibly related; UR = unrelated

*Each participant is counted only once at the highest level of severity for the event. This table represents severity of all adverse events sorted in descending order of incidence as shown above; or adverse events related to the intervention as judged by the investigator; or treatment emergent event.*

*AEs determined from both nasal exam or lab results findings (noted as ‘nasal exam’ and ‘labs’).*
